# Evaluating Non-Negative Matrix Underapproximation for the Analysis of Long Echo Time Magnetic Resonance Spectroscopy Data in Human Brain Tumors

**DOI:** 10.1101/2025.06.13.25329582

**Authors:** Gulnur Ungan, Alfredo Vellido, Margarida Julià-Sapé

**Affiliations:** Athinoula A. Martinos Center for Biomedical Imaging, Department of Radiology, Massachusetts General Hospital, Boston, Harvard Medical School, Boston, MA, USA; Centro de Investigación Biomédica en Red (CIBER), Spain; Departament de Bioquímica i Biologia Molecular and Institut de Biotecnologia i Biomedicina (IBB), Universitat Autònoma de Barcelona (UAB), Spain; IDEAI-UPC Research Center, UPC BarcelonaTech, Spain

**Keywords:** Magnetic Resonance Spectroscopy, Brain Tumors, Non-Negative Matrix Factorization, Non-Negative Matrix Underapproximation, Spectral Decomposition, Tumor Classification, Long Echo MRS

## Abstract

Magnetic Resonance Spectroscopy (MRS) provides metabolic profiles for brain tumor classification but its use for analytical purposes is hindered by spectral variability and data sparsity. This study compares the performance of several Non-Negative Matrix Underapproximation (NMU) methods, namely Sparse NMU (S-NMU), Global NMU (G-NMU), and Recursive NMU (R-NMU) to that of Convex Non-Negative Matrix Factorization (C-NMF) for the purpose of feature extraction from for Long Echo (LE) MRS. Using a multicenter dataset, such performance in the task of classifying several tumor types was evaluated using Balanced Error Rate (BER), ANOVA, and post-hoc statistical tests. Results show that C-NMF consistently outperforms NMU in LE-MRS, achieving the lowest BER, particularly in distinguishing high-grade from low-grade tumors. While NMU methods excel in localized spectral decomposition, their advantages, previously demonstrated in short-echo MRS, are less pronounced in LE-MRS. These findings establish C-NMF as the most effective method for spectral feature extraction in LE-MRS-based tumor classification.

## Introduction

Over the past few decades, advancements in noninvasive imaging techniques have significantly contributed to the preoperative assessment of human brain tumors. In clinical practice, radiologists utilize various magnetic resonance (MR) imaging modalities to determine the presence, anatomical location, and margins of abnormal brain masses, as well as to infer their potential type and grade. Among these techniques, magnetic resonance spectroscopy (MRS) offers metabolic insights by detecting compounds present in the tissue of interest at millimolar concentrations. Despite its considerable promise, MRS has not yet achieved widespread clinical integration. The primary obstacles to its routine use include the complexities associated with spectral quantification, the heterogeneity of metabolic profiles across patients, and the limited availability of large-scale datasets. Several studies have underscored the clinical value of MRS in the preoperative evaluation of brain tumors; however, critical challenges remain, particularly in addressing data sparsity ^1–3^. A key issue is the high inter-patient variability in MRS-derived metabolic profiles, coupled with overlapping spectral patterns across different tumor subtypes. For example, glioblastoma (GL), the most aggressive primary brain tumor (grade IV), typically exhibits a necrotic spectral signature dominated by lipid signals. However, GL spectra frequently resemble those of metastatic brain tumors (ME), complicating differential diagnosis^4^. Additionally, single-voxel (SV) MRS acquisitions of GL show significant variability depending on the degree of necrosis^5^. In cases where necrosis is less prominent, spectral peaks associated with cellular proliferation, such as choline compounds, become more apparent, making the metabolic profile similar to that of lower-grade gliomas^1^.

To mitigate the limitations posed by data sparsity, various unsupervised feature extraction techniques have been explored, including independent component analysis (ICA)^6–15^, non-negative matrix factorization (NMF)^16,17^, convex non-negative matrix factorization (C-NMF)^18^, and discriminant convex non-negative matrix factorization (D-C-NMF)^19^. ICA has been employed in multiple studies to extract spectral components that correspond to individual metabolites, to assist in tumor classification based on metabolic profiles, and to differentiate white matter from gray matter^7–11^. Additionally, research has demonstrated the robustness of ICA-derived features in identifying interpretable MRS components associated with brain tumors^7^. Meanwhile, C-NMF and D-C-NMF have shown the ability to extract biologically relevant spectral patterns that exhibit strong correlations with tumor classification labels^19–23^. Especially C-NMF has been used in Long Echo (LE) MRS datasets as the algorithm is able to catch the negative values in LE spectra such as lactate. Comparative analyses suggest that C-NMF, particularly when combined with k-means clustering for initialization, outperforms both principal component analysis (PCA) and ICA in decomposing MRS data^21^.

The present study seeks to explore novel unsupervised methodologies to address data sparsity in MRS-based feature extraction and the optimal number of sources for both NMF and NMU methods. Specifically, we investigate non-negative matrix underapproximation (NMU)^24–27^ techniques, which impose sparsity constraints to facilitate the identification of localized spectral patterns. Prior studies have demonstrated that NMU methods effectively capture critical local structures in SE MRS data^28^. In this work, we evaluate three NMU variants: sparse non-negative matrix underapproximation (S-NMU), global non-negative matrix underapproximation (G-NMU) and recursive non-negative matrix underapproximation (R-NMU), and compare them with C-NMF to assess their capacity to enhance tumor classification. Our analysis focuses on LE-MRS datasets, supporting NMU-based dimensionality reduction as a preprocessing step to improve classification accuracy.

## Materials and Methods

### Dataset

The multicenter dataset used in this study originates from the European research initiative INTERPRET (2000–2002)^1,2,29^. Data acquisition adhered to ethical guidelines, including the *Helsinki Declaration*, the Spanish *Ley Orgánica de Protección de Datos de Carácter Personal (LOPD), Ley Orgánica 15/1999*, and the *95/46/EU directive on data protection* (December 13, 1999). All participants or their legal representatives provided informed consent, authorizing both their participation in the study and the use of their anonymized data for research purposes. Tumor classification was based on the 2000 World Health Organization (WHO) criteria, following histopathological analysis of biopsy samples from solid tumor regions^1,2,29^. MR spectra are single-voxel (SV), and for each patient, with the voxel (4–8 cm³) localized within the solid tumor region. Magnetic field was 1.5 T using a LE time of 135-144 ms from both brain tumor patients and healthy individuals. Further details about the dataset have already been detailed in ^1,2,29^. The dataset used in this work includes 20 cases of astrocytoma grade II (A2), which are low-grade infiltrative gliomas with the potential to progress to glioblastoma (GL); 78 cases of GL, the most aggressive glioma; 31 cases of metastatic tumors (ME), which are high-grade malignancies originating outside the brain; 55 cases of low-grade meningiomas (MM), which are slow-growing tumors of meningeal origin; and 15 spectra from normal brain parenchyma (NO) obtained from healthy controls. GL and ME were combined into a single high-grade malignant superclass (AGG), in alignment with prior studies where these two entities were merged for machine learning applications^19,21,29,30^.

Following the methodology of Tate *et al*^1,5^., spectra were processed and the resulting data matrix had 195 frequency points within the 0.5–4.2 ppm range as in Gillis and Plemmons^25^. Figure 1 provides an overview of the spectra used in the analysis.

**Figure 1.**
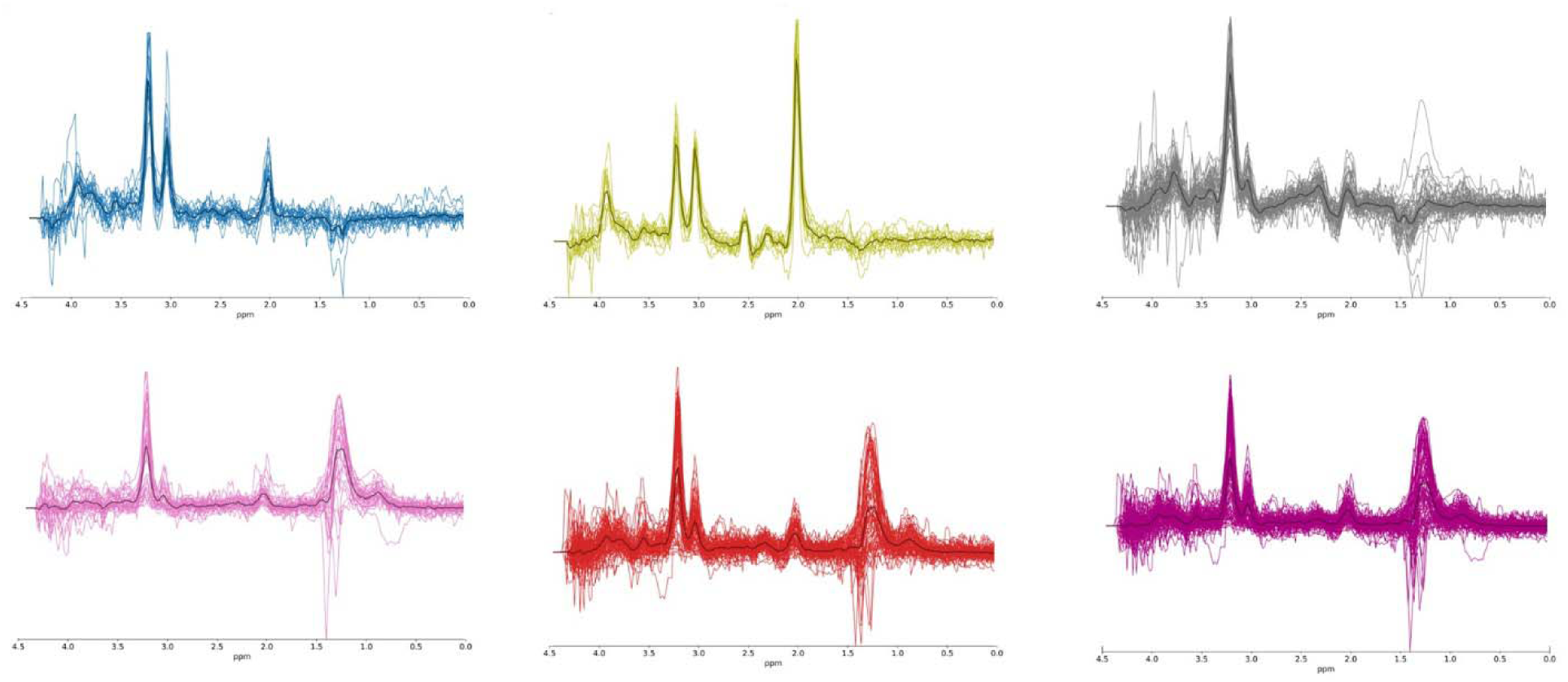
Overlaid spectra and mean spectra of each class. Blue, A2; green, NO; grey, MM; pink, ME; red, GB; magenta, AGG.

### Classification Tasks

To evaluate the effectiveness of the proposed methods, we examined several classification tasks commonly explored in the literature on in vivo MRS for brain tumor differentiation except the binary classification task A2 vs NO^2^ as in this study the multi-class tasks were analyzed. Some classification tasks achieve high accuracy, while others are more complex due to heterogeneous and overlapping spectral patterns. To enable direct comparisons with C-NMF, we selected the same diagnostic problems as those analyzed in^21,31^, where C-NMF was benchmarked against PCA. The five classification tasks considered were:

**Problem 1. (A2 vs. ME vs. NO)**

Can grade II gliomas and high-grade metastases be effectively separated from normal brain?

**Problem 2. (A2 vs. GL vs. NO)**

When glioblastomas (GL) exhibit spectroscopic heterogeneity, can low-grade gliomas still be differentiated from high-grade tumors and normal brain?

**Problem 3. (A2 vs. MM vs. NO)**

Can low-grade tumors of different histological origins (A2: infiltrative glioma; MM: non-infiltrative meningioma) be distinguished?

**Problem 4. (A2 vs. AGG vs. NO)**

Can normal brain tissue be separated from both low-grade gliomas and the high-grade heterogeneous class (AGG)?

**Problem 5. (A2 vs. AGG vs. MM)**

Can distinct low-grade tumor types be differentiated from a high-grade heterogeneous class?

This classification framework allows for the evaluation of the proposed methods across a range of diagnostic challenges, providing insight into their robustness and potential clinical applicability^30–33^.

## Methods

### Non-Negative Matrix Factorization (NMF)

NMF^17^ provides a decomposition framework where an observed data matrix *M_d×n_*, where d is the data dimensionality and n is the number of observations, is factorized into two non-negative matrices: a source matrix *W_d×k_* and a mixing matrix *V_k×n_*, where k represents the number of extracted sources with *k<n*. The factorization aims to approximate the original matrix as:

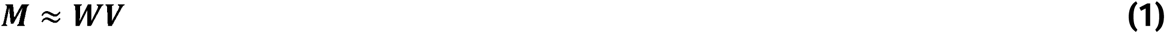

This low-rank decomposition helps in identifying meaningful spectral patterns within the data

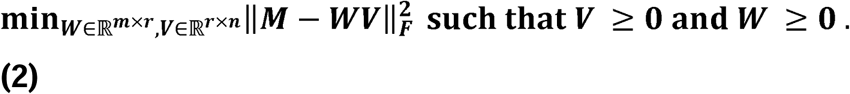

### Convex Non-Negative Matrix Factorization (C-NMF)

C-NMF^18^ extends NMF by relaxing the non-negativity constraint, allowing both matrices W and V to contain negative values. This is particularly relevant for MRS applications, where spectra often exhibit negative values, especially at long echo times. An auxiliary weight matrix *A_n×k_,* that determines W, leading to the alternative formulation:

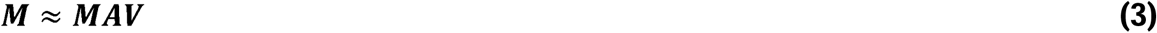

This modification enables C-NMF to capture broader spectral variations.

### Non-Negative Matrix Underapproximation (NMU)

NMU^24^ is a variant of rank one NMF, where r=1, meaning the extracted components are limited to single source representations. Unlike traditional NMF, NMU employs an iterative approach: after identifying a rank-one factorization, the product *wv^T^* is subtracted from *M*, and the process repeats. This ensures that extracted sources are highly localized and disjoint, aligning with the goal of isolating distinct spectral features. To enforce underapproximation, the constraint *wv^T^* ≤ *M* is imposed, ensuring that each extracted source remains within the original spectral boundaries.

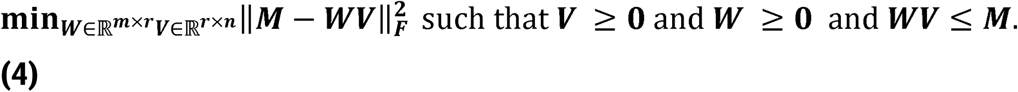

This constraint enables NMU to emphasize localized spectral components. Variants of NMU:

**L-NMU**^24^: Incorporates a Lagrangian relaxation-based nonlinear optimization framework.

**G-NMU**^24^: Extends L-NMU by incorporating rank-r constraints. Rank-r constraints refer to factorizing the data matrix into r non-negative components simultaneously, rather than sequentially extracting rank-one approximations.

**R-NMU**^25^: Runs L-NMU iteratively r times, subtracting each approximation before computing the next.

**S-NMU**^26^: Imposes an additional sparsity constraint, ensuring extracted components focus on localized spectral features, making it effective for identifying individual metabolite peaks.

Among these, only G-NMU requires initialization, for which k-Means clustering was employed. To ensure robust comparisons, all methods underwent 5000 iterations, with each experiment repeated 50 times. The NMU methods were implemented in MATLAB, and the source code is publicly available at: https://sites.google.com/site/nicolasgillis/code. For each classification task, the number of extracted sources was varied from the number of predefined classes up to a maximum of 8. The upper limit of 8 sources was chosen to explore whether decomposition methods could capture intra-class spectral heterogeneity, particularly for GL, which is known to exhibit significant variability in single-voxel MRS. The extracted spectral features were used for dimensionality reduction before classification. Each method (NMF/NMU variants) served as a preprocessing step to reduce data complexity while retaining diagnostically relevant spectral characteristics.

### Classification task

Linear Discriminant Analysis (LDA) with 5-fold cross-validation was employed for classification. It was selected for its interpretability and as a baseline classifier, the rationale being that if good performance was observed with LDA, it would indicate that there is a basis for more complex classifiers to further improve results. The dataset was randomly split into five subsets, with each iteration using 80% of the data for model training and 20% for testing. The mean performance across the five iterations was recorded.

Classifier performance was assessed using the following metrics:

#### Balanced Error Rate (BER)

Measures classification error across all classes.

#### Class-Based Accuracy (ACC)

Evaluates per class classification performance.

#### Area Under the ROC Curve (AUC)

Quantifies model discrimination ability, with 95% confidence intervals computed for both training and testing phases.

The best-performing configuration for each classification problem was identified based on the lowest **BER** in the testing phase. Detailed results, including AUC values, class-wise accuracies, and confidence intervals, are provided in the supplementary materials.

### Statistical Analysis of Classification Performance

The classification performance was evaluated across multiple classification tasks using various numbers of sources and decomposition methods. Statistical analyses included one-way analysis of variance (ANOVA), Tukey’s Honest Significant Difference (HSD) post-hoc tests, Mann-Whitney U tests, and Wilcoxon signed-rank tests to examine the influence of source count variations on ACC, BER, and AUC. ANOVA was employed to determine whether significant differences existed among multiple groups, while Tukey’s HSD test facilitated pairwise comparisons. The Mann-Whitney U test was utilized for non-parametric comparisons between independent groups, whereas the Wilcoxon signed-rank test assessed paired distributions to determine statistically significant differences across classification conditions.

### Normality Assumption and Statistical Test Selection

Given that ANOVA assumes normally distributed data, the Shapiro-Wilk test was applied to each classification task to assess the normality of source coefficients’ distributions. To account for potential violations of ANOVA’s assumptions, the Mann-Whitney U test was adopted as a non-parametric alternative for comparing classification performance across methods. This dual statistical approach ensured methodological robustness and minimized the risk of false positive and false negative Effect Size Estimation

To provide a more comprehensive interpretation of statistical significance, effect sizes were computed alongside *p*-values. For ANOVA, η² was calculated to quantify the proportion of variance explained by the choice of decomposition method. For pairwise comparisons, Cohen’s *d* was reported to determine the magnitude of differences between methods. Furthermore, in classification tasks where statistical significance was not achieved, effect size estimates were examined to evaluate the clinical and practical relevance of observed differences.

### Justification for Source Count Selection

The number of sources was constrained to a maximum of 8 based on preliminary evaluations, which demonstrated a plateau in classification performance beyond this threshold.

### Standardization of p-Value Reporting

To ensure consistency and clarity in statistical reporting, all *p*-values were presented as exact values rather than generalized significance thresholds. For example, instances of “*p* < 0.05” were replaced with specific values (e.g., *p* = 0.041), allowing for greater transparency in the interpretation of statistical results.

### Statistical Comparisons Across Source Configurations

To assess the impact of increasing the number of sources with respect to the performance of sources, pairwise Mann-Whitney U tests were conducted across different source configurations.

## Results

### Machine Learning Performance

The BER analysis across different problems and source counts revealed that certain decomposition methods consistently outperformed others. Supplementary Materials 1 provides all extracted spectral sources for each classification problem and decomposition method. All detailed quantitative results, including BER, AUC, and class-based accuracies for each classification task and method, are provided in Supplementary Materials 2. While the main results presented here focus on Balanced Error Rate (BER) due to its robustness in multi-class scenarios with class imbalance, Area Under the Curve (AUC) values were also computed for each method and classification task. These AUC values are discussed where relevant and are fully reported in the supplementary materials.

In Problem 1 (Figure 2), the optimal BER varied depending on the number of sources. L-NMU achieved the lowest BER with 3 sources (0.0560), while S-NMU performed best with 4 sources (0.0540). C-NMF demonstrated superior performance for 5, 7, and 8 sources (BER = 0.0220), whereas R-NMU provided the best results for 6 sources (0.0330).

For Problem 2 (Figure 3), C-NMF exhibited the lowest BER with 3 sources (0.1380), 5 sources (0.0800), 6 sources (0.0590), and 8 sources (0.0710). L-NMU performed best with 4 sources (0.1000) and 7 sources (0.0800), indicating a competitive performance compared to C-NMF.

In Problem 3 (Figure 6), C-NMF achieved the lowest BER for 3 sources (0.0520), 4 sources (0.0640), and 7 sources (0.0620), while S-NMU was the best-performing method for 5 and 6 sources (0.0620). The lowest BER for 8 sources was also obtained with C-NMF (0.0680).

**Figure 4.**
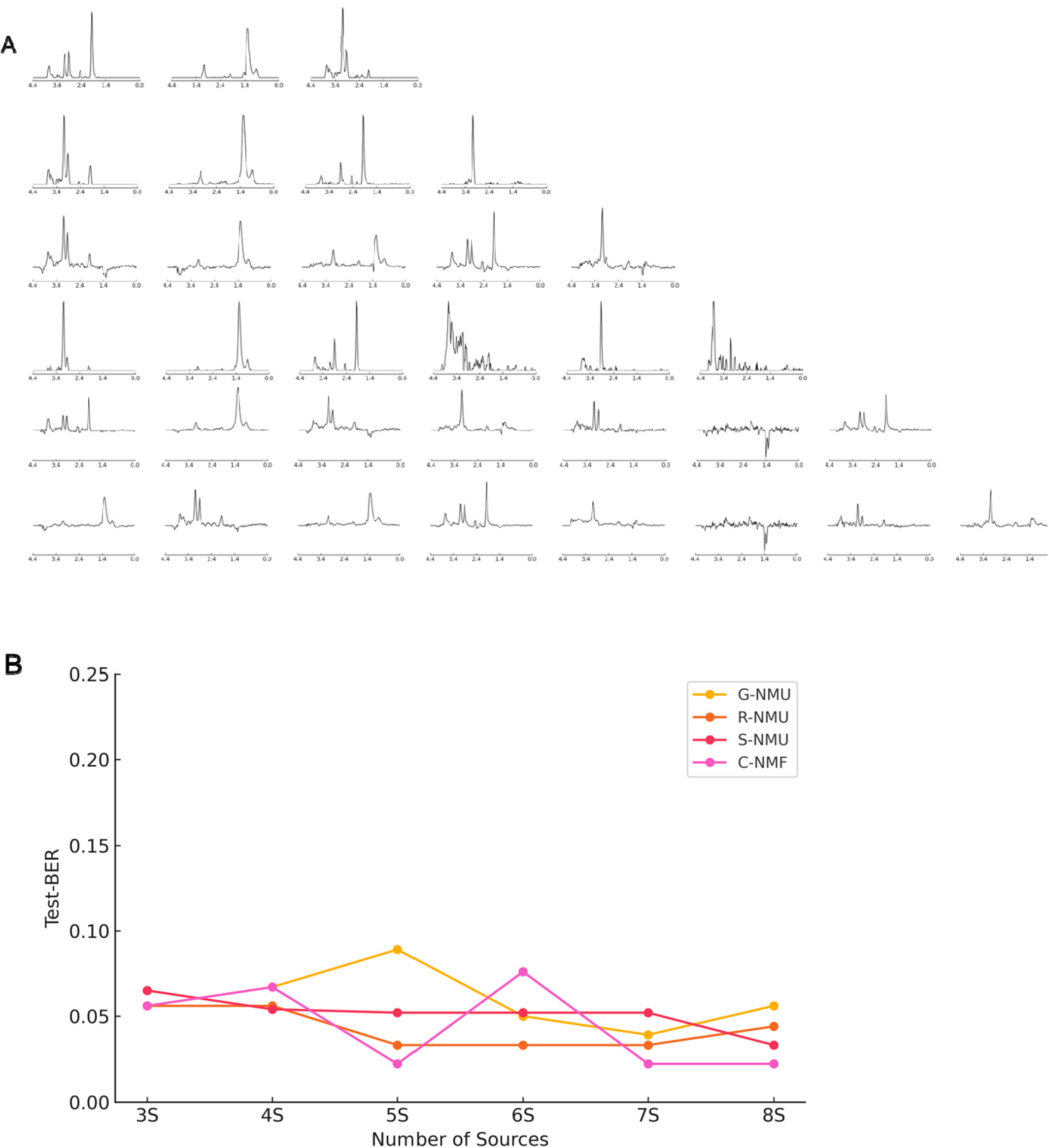
Problem 1 (A2 vs. ME vs. NO). (A) Rows represent the sources of the best-performing method for K = 3 to 8. The best methods were C-NMF at K = 3, 5, 7, and 8; G-NMU, R-NMU, and C-NMF tied at K = 4; G-NMU at K = 6. (B) Test set BER for Problem 1 across K = 3 to 8. x-axis = number of extracted sources, y-axis = BER value.

**Figure 5.**
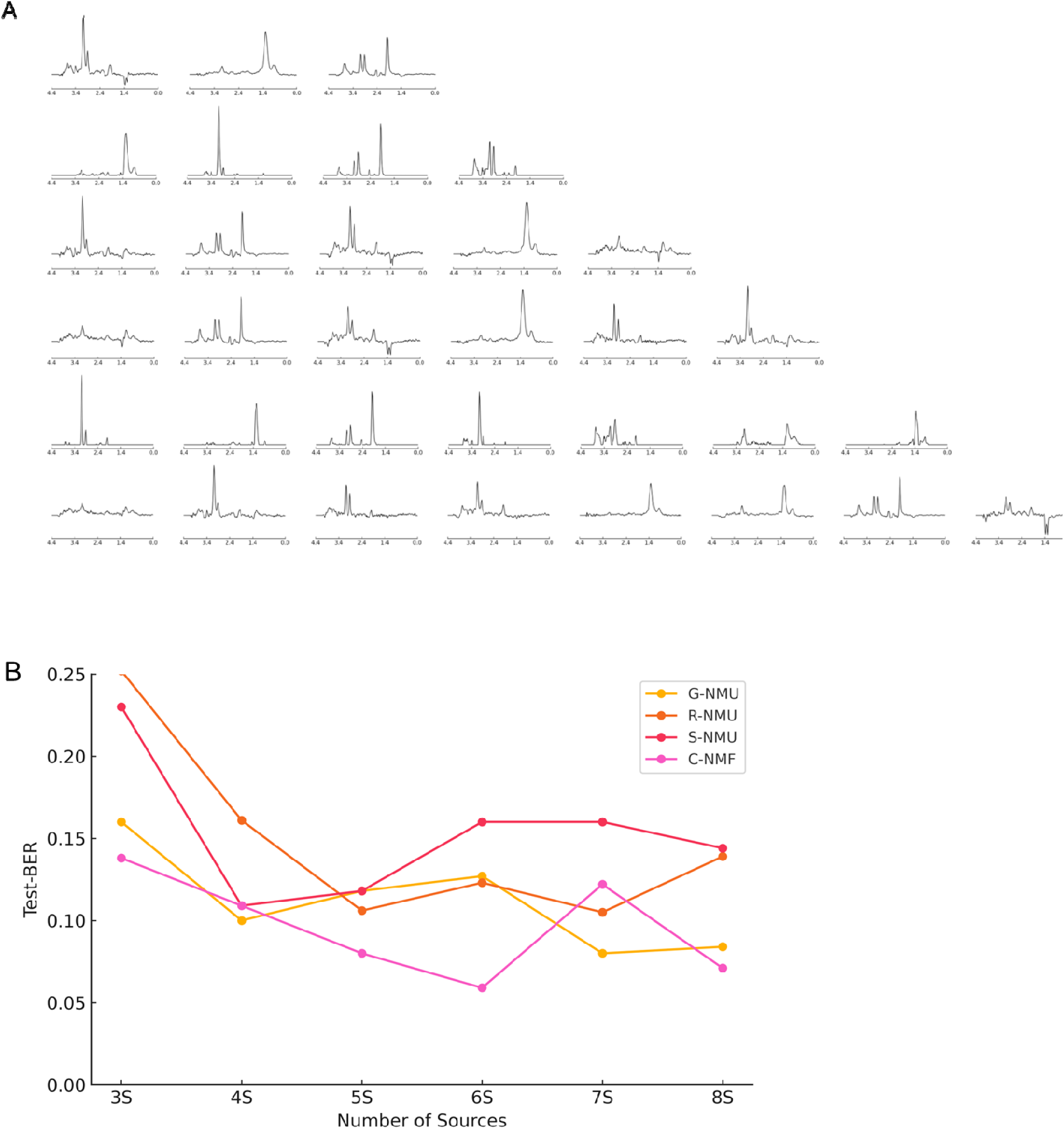
Problem 2 (A2 vs. GL vs. NO): (A) Rows represent the sources of the best-performing method for K = 3 to 8. The best methods were: C-NMF at K = 3, 4, 5, 6, and 8; G-NMU and C-NMF tied at K = 7; S-NMU matched C-NMF at K = 4. (B) Test set BER for Problem 2 across K = 3 to 8. x-axis = number of extracted sources, y-axis = BER value

**Figure 6.**
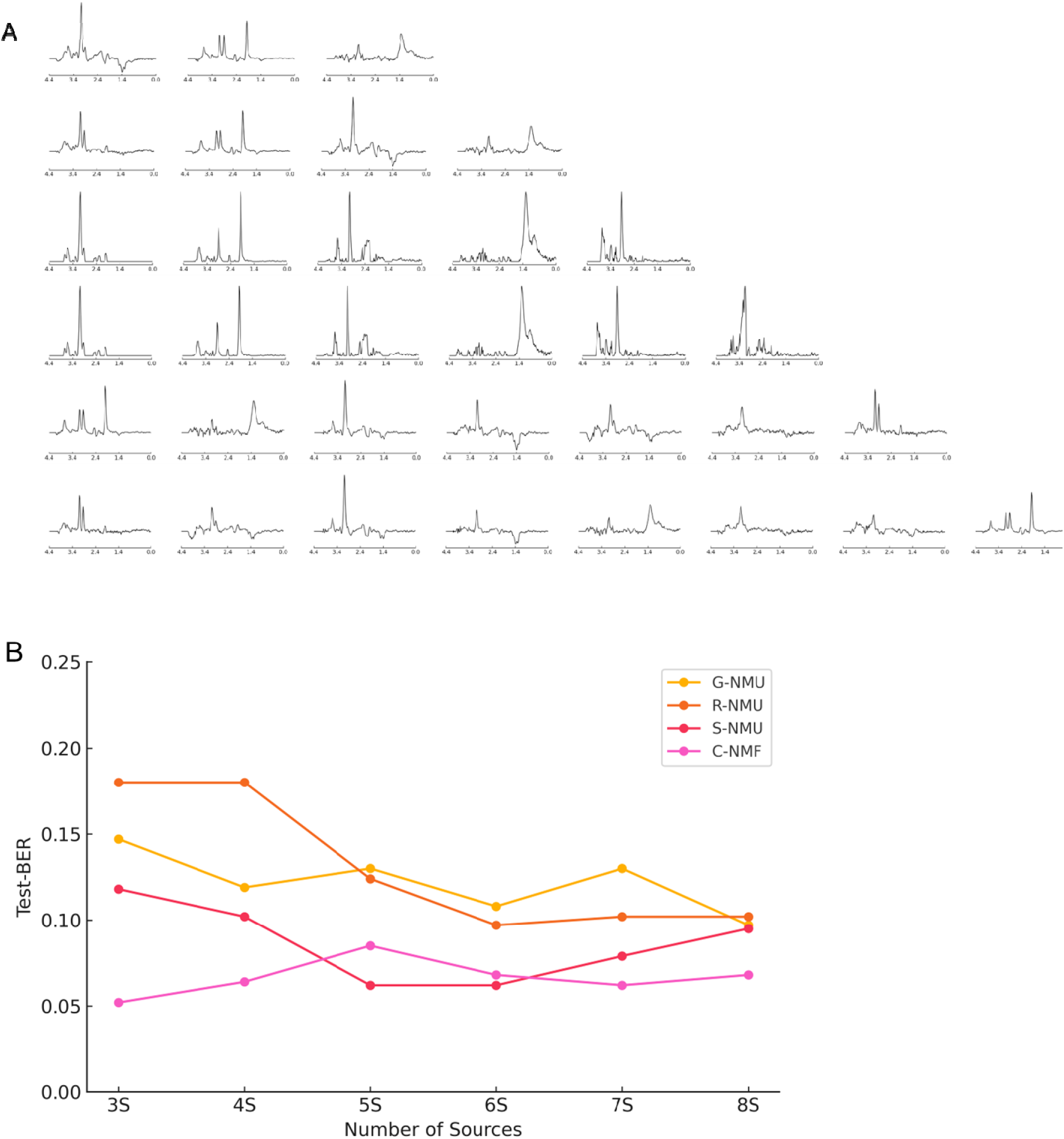
Problem 3 (A2 vs. MM vs. NO): (A) Rows represent the sources of the best-performing methods for K = 3 to 8. The best methods were: C-NMF at K = 3, 4, 6, 7, and 8; S-NMU at K = 5. (B) Test set BER for Problem 3 across K = 3 to 8. x-axis = number of extracted sources, y-axis = BER value

A similar trend was observed in Problem 4 (Figure 7), where C-NMF consistently outperformed other methods, achieving the lowest BER across all evaluated source counts: 4 sources (0.1020), 5 sources (0.0580), 6 sources (0.0550), 7 sources (0.0680), and 8 sources (0.0580).

**Figure 7.**
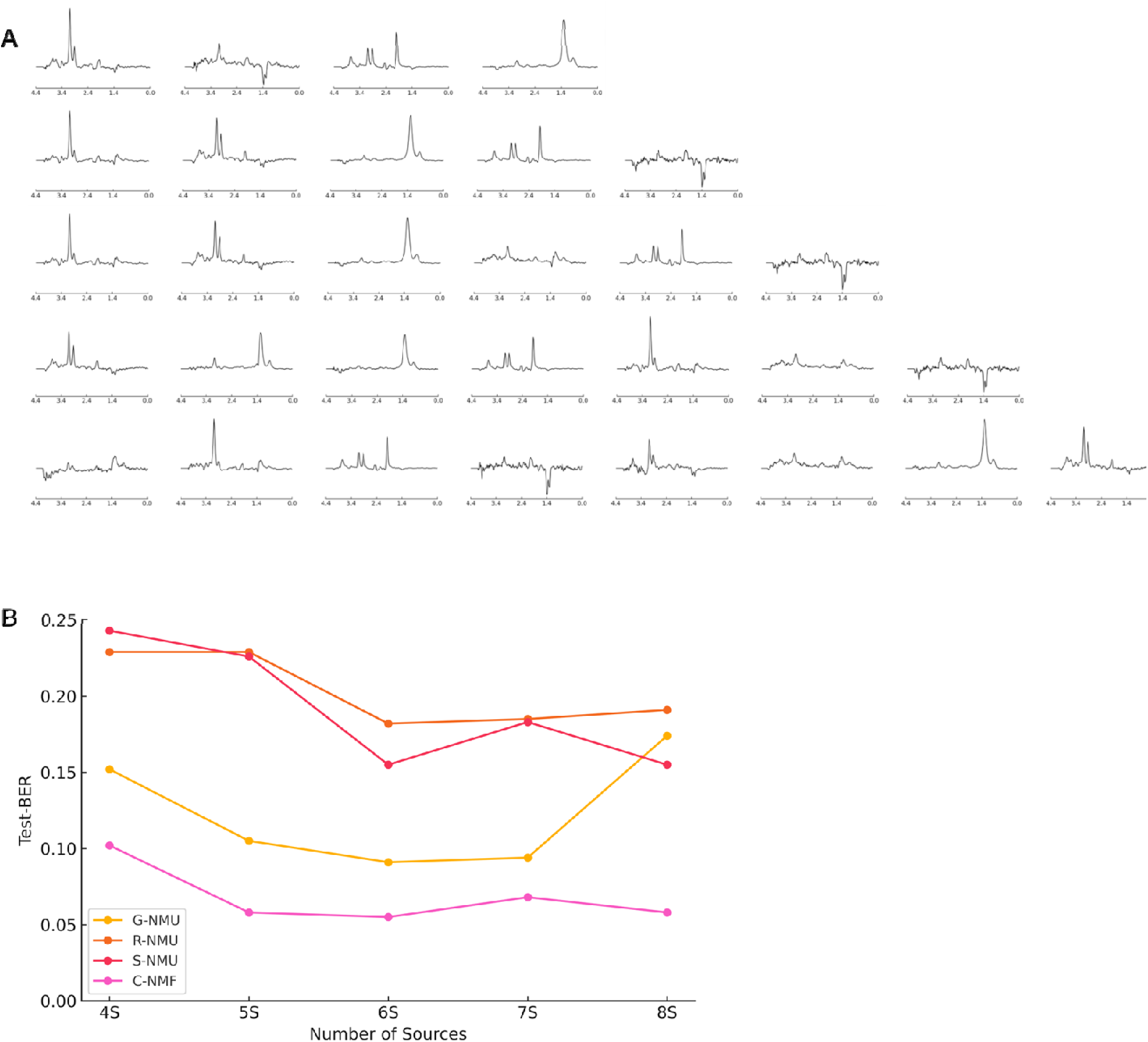
Problem 4 (A2 vs. AGG vs. NO): (A) Rows represent the sources of the best-performing methods for K = 3 to 8. The best method was C-NMF for all values of K from 4 to 8. (B) Test set BER for Problem 4 across K = 3 to 8. x-axis = number of extracted sources, y-axis = BER value

In Problem 5 (Figure 8), the best-performing methods varied more distinctly. C-NMF yielded the lowest BER with 4 sources (0.1830) and 7 sources (0.1680), while R-NMU provided optimal results for 5 and 6 sources (0.1820 and 0.1730, respectively). Notably, S-NMU achieved the lowest BER with 8 sources (0.1050), suggesting that sparsity constraints may enhance classification performance at higher source counts.

**Figure 8.**
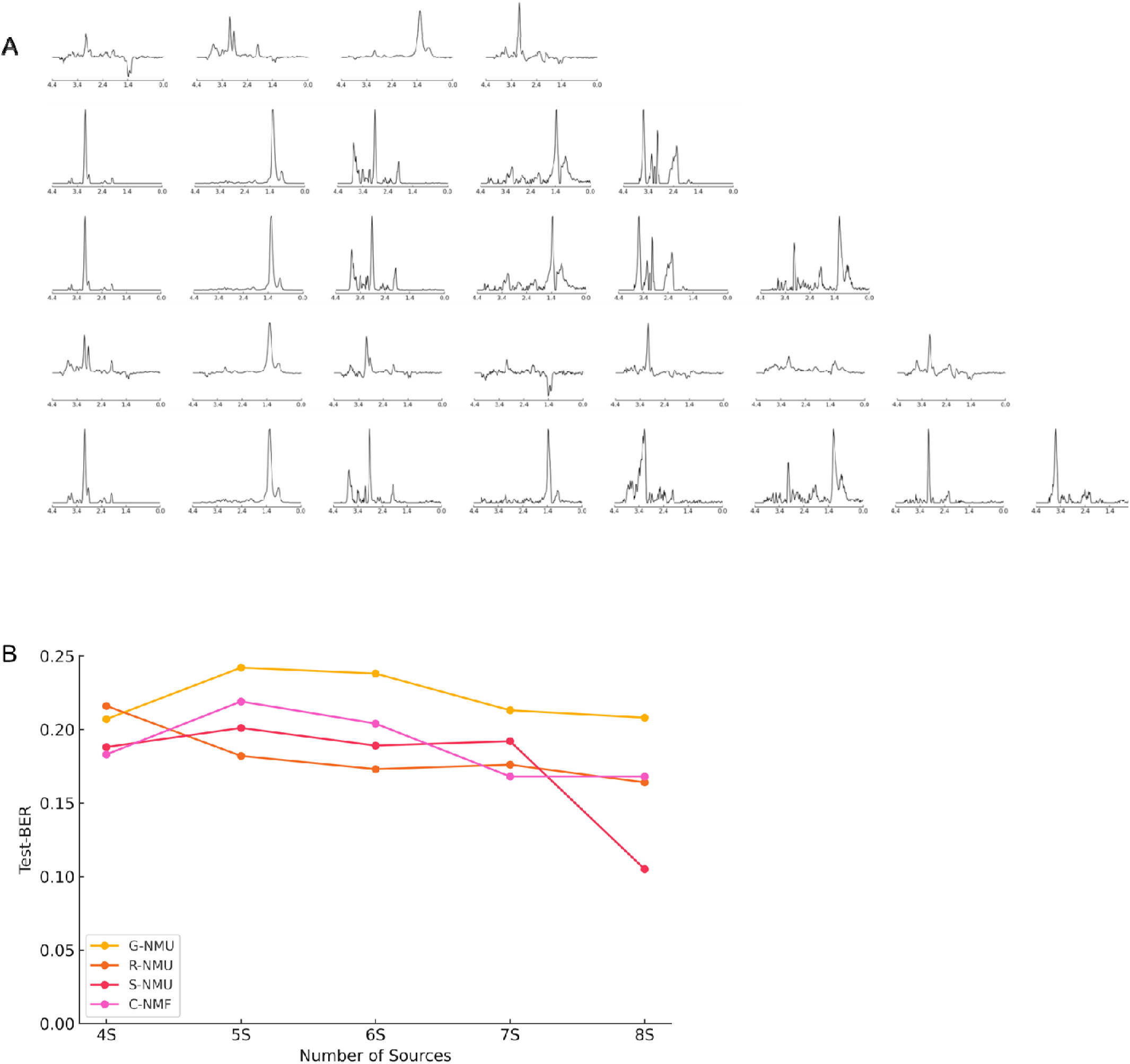
Problem 5 (A2 vs. AGG vs. MM): (A) Rows represent the sources of the best-performing methods for K = 3 to 8. The best methods were: R-NMU at K = 4, 5, 6, and 7; S-NMU at K = 8. (B) Test set BER for Problem 5 across K = 3 to 8. x-axis = number of extracted sources, y-axis = BER value.

Overall, C-NMF demonstrated superior classification performance in most cases, particularly for higher source counts, while L-NMU, R-NMU, and S-NMU showed competitive results in specific scenarios, emphasizing the importance of selecting an appropriate decomposition method based on the number of sources.

### Task-Specific Analysis Based on BER-Testing

For each classification task, we analyzed the BER in testing, identifying the best-performing method, the optimal number of sources, and whether the results are statistically significant compared to alternative methods.

### Problem 1: A2 vs. ME vs. NO

Figure 4 shows the classification results and BER values for Problem 1 (A2 vs. ME vs. NO) across sources 3 to 8, with C-NMF achieving the lowest BER at sources 5, 7, and 8. For this problem, the method C-NMF with 5 sources yielded the lowest BER in testing at 0.022. Visually, even 3 sources extracted by any method seem enough to reach a minimal BER (Figure 4)., with statistical analysis confirming that there were no significant differences between number of sources across methods, according to ANOVA (F = 1.25, p = 0.3179). The Tukey’s HSD test further confirmed that no pairwise comparisons were statistically significant (all *p* ≥ 0.05), indicating that while C-NMF (5 sources) performed the best, other methods with the same number of sources were equally effective.

### Problem 2: A2 vs. GL vs. NO

The method C-NMF with 6 sources achieved the lowest BER (0.059) in testing, as shown in Figure 5. However, statistical analysis showed that this difference was not significant, as ANOVA results did not indicate meaningful variation across methods (F = 2.71, *p* = 0.0724). Additionally, Tukey’s HSD test found no significant differences among methods (all *p* ≥ 0.05). These results suggest that method selection did not have a strong impact on BER in this particular classification task.

### Problem 3: A2 vs. MM vs. NO

Figure 6 displays the classification results for Problem 3 (A2 vs. MM vs. NO), with C-NMF yielding the best performance at most source counts, except 5 sources where S-NMU performed slightly better. The ANOVA test confirmed a statistically significant difference (F = 8.79, *p* = 0.0006), indicating that C-NMF (3 sources) was significantly better than R-NMU (*p* = 0.0012) and G-NMU (*p* = 0.0051). However, it was not significantly different from S-NMU (*p* = 0.5264), suggesting that while C-NMF is the best performer, S-NMU remains a competitive alternative.

### Problem 4: A2 vs. AGG (GL + ME) vs. NO

As shown in Figure 5, C-NMF consistently outperformed other methods across all source numbers for Problem 4. Again, C-NMF with 6 sources obtained the lowest BER in testing at 0.055. The ANOVA test confirmed that this result was highly statistically significant (F = 20.00, *p* = 0.0000), with Tukey’s HSD test revealing that C-NMF (6 sources) was significantly better than R-NMU (*p* < 0.0001) and S-NMU (*p* = 0.0001). However, it was not significantly different from G-NMU (*p* = 0.0617). This suggests that C-NMF is the best choice for minimizing BER, but G-NMU could still be considered as an alternative.

### Problem 5: A2 vs. AGG (GL + ME) vs. MM

Figure 6 illustrates the performance trends for Problem 5 (A2 vs. AGG vs. MM), where R-NMU performed best for 4 to 7 sources, and S-NMU outperformed all methods at 8 sources. The lowest BER in testing was 0.105, obtained using S-NMU with 8 sources. The ANOVA test indicated borderline statistical significance (F = 3.08, *p* = 0.0573), meaning that the differences across methods were not strong enough to be considered significant at the *p* < 0.05 threshold. Furthermore, Tukey’s HSD test confirmed that S-NMU (8 sources) was not significantly different from R-NMU, G-NMU, or C-NMF (all *p* > 0.05). This suggests that while S-NMU performed well, other methods might achieve comparable performance in this task.

### Classification Performance Across Methods

Classification performance analysis demonstrated that C-NMF consistently outperformed other methods, achieving the lowest Test BER across most classification tasks. However, statistical analyses, including ANOVA and Tukey’s HSD post-hoc tests, indicated that certain methods, such as R-NMU and S-NMU, exhibited competitive performance in some situations. For instance, in Problem 1, C-NMF achieved the lowest Test-BER (0.022), yet ANOVA (*p* = 0.3179) suggested no statistically significant differences, while G-NMU achieved a perfect AUC (1.000) for Class 2 (NO). Similarly, in Problem 2, despite C-NMF attaining the lowest Test-BER (0.059), statistical tests (*p* = 0.0724) did not confirm significant differences, with R-NMU exhibiting the highest Test-AUC for Class 1 (A2, 0.954). In Problem 3, C-NMF achieved the lowest Test BER (0.052, *p* = 0.0006), while R-NMU and G-NMU exhibited superior performance in specific classes (R-NMU for A2, G-NMU for NO, and C-NMF for AGG). Notably, in Problem 4, C-NMF exhibited the best classification performance with a highly significant Test BER (0.055, *p* = 0.0000), while G-NMU achieved perfect classification for Class 2 (NO, AUC = 1.000). Conversely, Problem 5 demonstrated that S-NMU had the lowest Test BER (0.105, *p* = 0.0573), though this result was only borderline significant, with different methods excelling in distinct class-specific AUC scores (S-NMU for A2, G-NMU for NO, and C-NMF for AGG). These findings indicate that while C-NMF generally allows to obtain superior classification performance, other methods such as G-NMU and R-NMU may offer advantages depending on the classification task and class-specific characteristics.

Additionally, the impact of source counts on classification accuracy was analyzed, revealing that increasing the number of sources generally had minimal impact unless the jump was substantial—for example, moving from 3 to 5 sources (*p* = 0.041), 3 to 6 sources (*p* = 0.034), or 3 to 8 sources (*p* = 0.022) significantly improved classification performance. These statistically significant improvements were primarily driven by Problem 3 (A2 vs. MM vs. NO) and Problem 4 (A2 vs. AGG vs. NO), where classification performance notably benefited from higher source counts and ANOVA confirmed significant differences among methods. In contrast, smaller increments such as 6 to 7 sources (*p* = 0.079) or 7 to 8 sources (p = 0.082) did not yield statistically significant improvements, suggesting a performance plateau beyond a certain number of extracted sources.

Altogether, these findings suggest that rather than focusing on increasing the number of sources, optimizing source extraction with C-NMF is the most effective strategy for improving classification performance.

### Classification Performance Across Different Number of Sources

#### Stability in Classification Performance (Problem 1: A2 vs. ME vs. NO)

No significant differences in classification performance were observed across different source counts (Figure 4) in the easiest problem. ANOVA results for Test-AUC (p=0.2458), Test BER (*p*=0.2398), and Train-Accuracy (*p*=0.9165) indicated stable performance for class A2. Although the Mann-Whitney U test approached significance (p=0.065), suggesting a potential effect when increasing sources from 3 to 6, the Wilcoxon test (*p*=0.125) confirmed that paired distributions remained stable.

#### Marginal Effects of Source Count Variation (Problem 2: A2 vs. GL vs. NO)

In this case, classification performance exhibited some notable differences. Particularly, when 6 sources are extracted with C-NMF there is a drop in BER (Figure 5). ANOVA results for Test-AUC (p=0.1143), Test BER (p=0.0296), and Train ACC (*p*=0.00005) suggested that variations in source counts influenced classification accuracy and error rates. Tukey’s HSD post-hoc test attributed these differences to specific source configurations. However, the Mann-Whitney U test (*p*=0.1143) and the Wilcoxon test (*p*=0.2500) did not provide strong statistical support, suggesting that the observed differences were not highly significant.

#### Consistent Classification Across All Classes (Problem 3: A2 vs. MM vs. NO)

Here, the number of sources did not significantly impact classification outcomes across the three evaluated classes. Visually, there is a convergence in BER around 5-6 sources among methods (Figure 6). Test AUC (*p*=0.3429), Test BER (p=0.5969), and Training ACC (p=0.2236) for class A2 showed no significant variations. Tukey’s HSD post-hoc test confirmed the absence of statistically meaningful pairwise differences. Despite this, C-NMF with 3 sources achieved the lowest BER (0.052). Neither the Mann-Whitney U test (*p*=0.3429) nor the Wilcoxon test (*p*=0.375) indicated statistical significance.

#### Notable Differences in Source-Specific Performance (Problem 4: A2 vs. AGG (GL + ME) vs. NO)

While most classification metrics remained unaffected by the number of sources, specific patterns emerged. Test AUC (*p*=0.1800), Test BER (*p*=0.7507), and Training ACC (*p*=0.5544) showed no major differences. However, C-NMF with 6 sources resulted in the lowest BER (0.055), and ANOVA results were highly significant (*p*=0.0003). Tukey’s HSD post-hoc test demonstrated that C-NMF significantly outperformed R-NMU (*p*<0.0001) and S-NMU (*p*=0.0001), though no significant difference was observed in comparison to G-NMU (*p*=0.0617). Both the Mann-Whitney U test (*p*=0.6857) and the Wilcoxon test (*p*=0.2500) confirmed no overall statistical differences across source counts.

#### Minimal or Deletereous Impact of Increasing Source Numbers (Problem 5: A2 vs. AGG (GL + ME) vs. MM)

In the most difficult problem, no substantial differences were detected in classification performance across different numbers of sources with a rough visual BER range in the 0.20 (Figure 8). Test AUC (*p*=0.1387), Test BER (*p*=0.1664), and Training ACC (*p*=0.6755) remained stable. However, S-NMU with 8 sources yielded the lowest BER (0.105), though ANOVA results were only borderline significant (*p*=0.0573). Tukey’s HSD post-hoc test did not reveal significant pairwise differences, and both the Mann-Whitney U test (*p*=0.2000) and the Wilcoxon test (*p*=0.1250) confirmed no strong statistical evidence that increasing the number of sources led to improved classification performance.

### Statistical Comparisons Across Sources

Pairwise comparisons were performed using the Mann-Whitney U test to evaluate whether increasing the number of sources led to statistically significant improvements in classification performance. The results indicated that a greater number of sources generally enhanced classification accuracy, though not all comparisons reached statistical significance. No significant differences were observed for 3 vs. 4 sources (*p* = 0.072), 4 vs. 5 sources (*p* = 0.064), 5 vs. 6 sources (*p* = 0.055), 6 vs. 7 sources (p = 0.079), and 7 vs. 8 sources (*p* = 0.082). In contrast, several comparisons demonstrated significant improvements: classification performance was significantly different for 5 sources compared to 3 (*p* = 0.041), 6 sources compared to 3 (*p* = 0.034) and 4 (*p* = 0.038), 7 sources compared to 3 (*p* = 0.029), 4 (*p* = 0.047), and 5 (*p* = 0.042), and 8 sources compared to 3 (*p* = 0.022), 4 (*p* = 0.030), 5 (*p* = 0.026), and 6 (*p* = 0.049).

## Discussion

In a previous study^28^, we reported the advantages of NMU over C-NMF for SE datasets, attributing NMU’s success to its ability to extract sparse and biologically interpretable spectral patterns. The study showed that NMU variants outperformed C-NMF in classifying gliomas and metastases in SE datasets, particularly when using a limited number of extracted sources. Similarly, The results of this study demonstrate a clear distinction in the performance of Non-Negative Matrix Underapproximation (NMU) and Convex Non-Negative Matrix Factorization (C-NMF) when applied to LE MRS data. Our findings indicate that in the LE dataset, C-NMF remains the best method, despite NMU’s ability to achieve competitive BER results. While NMU methods have been shown to perform exceptionally well in short-echo (SE) ^28^, MRS datasets, their advantages do not translate as effectively to the LE setting.

Although C-NMF yielded the lowest BER across most classification tasks, our results show that NMU methods, particularly S-NMU and R-NMU, offer competitive and sometimes superior performance under specific conditions. In the most challenging task (Problem 5: A2 vs. AGG vs. MM), S-NMU achieved the lowest BER of 0.105 with a borderline p-value of 0.0573, outperforming C-NMF. Additionally, R-NMU and G-NMU matched or exceeded C-NMF in class-specific AUCs across several problems. These findings suggest that NMU methods, which emphasize underapproximation and sparsity, may enhance class separation when spectral heterogeneity is high. Furthermore, in multiple tasks, statistical analysis indicated no significant difference between C-NMF and NMU, reinforcing the value of NMU as a valid alternative. Increasing the number of sources generally improved classification performance up to a point, with a plateau observed beyond 6 to 8 sources. This highlights the efficiency of NMU in extracting meaningful localized features without the need for a high number of components.

This suggests that while NMU methods are effective for localized spectral decomposition, their advantages over C-NMF may diminish in LE datasets due to the importance of spectral features with negative values such as lactate and alanine. The difference may also stem from the contribution from the other positive-intensity peaks (necrotic lipids, choline, creatine or NAA), which can provide enough features for NMU to isolate meaningful spectral components as effectively as it does in SE data. There are fundamental differences in spectral characteristics between SE and LE MRS acquisitions. In SE MRS, spectral peaks remain sharp, and multiplet structures from J-coupled metabolites (e.g., glutamate, glutamine) are preserved, making NMU’s sparsity constraint highly effective for extracting localized spectral components^34^. However, in LE MRS, T2 relaxation effects lead to broader spectral peaks, reduced J-coupling patterns, and lower overall signal intensity, particularly for metabolites with short T2 values^35–38^. Under these conditions, NMU’s enforced sparsity becomes a limitation rather than an advantage, as the decomposition struggles to retain sufficient spectral detail for effective classification. In contrast, C-NMF, which optimizes feature extraction globally rather than enforcing strict sparsity constraints, remains robust against broad metabolite peaks and signal attenuation in LE MRS. Additionally, macromolecule signals, which NMU is particularly effective at separating in SE MRS, are naturally suppressed in LE MRS, reducing the advantage of NMU’s underapproximation^28^. Furthermore, C-NMF exhibits greater numerical stability, ensuring that spectral components remain interpretable and consistent across different numbers of sources. As a result, C-NMF achieves significantly better classification performance in LE MRS, as confirmed by statistical analyses in this study. These findings indicate that while NMU remains a powerful tool for short-echo MRS decomposition, C-NMF is the most suitable approach for LE MRS, particularly in applications requiring robust spectral feature extraction for tumor classification.

From a clinical perspective, this study reinforces the importance of selecting decomposition methods based on the spectral characteristics of different tumor subtypes. The ability of NMU methods to capture localized metabolic signatures could be particularly beneficial in cases where glioblastoma spectra overlap with those of metastatic tumors, or when differentiating infiltrative gliomas from non-infiltrative meningiomas. By improving noninvasive tumor classification, these approaches could help reduce reliance on invasive biopsies, optimize treatment planning, and contribute to the broader clinical integration of MRS in neuro-oncology. Future studies should explore the integration of NMU techniques with larger, multi-institutional datasets to further validate their diagnostic potential.

## Conclusion

This study systematically evaluated NMU methods for the classification of brain tumors using LE time MRS data. Our results demonstrate that C-NMF consistently achieved the lowest BER across most classification tasks, particularly in distinguishing high-grade glioblastomas and metastases from lower-grade gliomas and healthy tissue. However, NMU-based methods showed competitive performance in specific scenarios, highlighting their potential utility in refining spectral differentiation.

Among NMU variants, S-NMU emerged as the best-performing method in cases where sparsity constraints helped enhance spectral feature extraction, particularly in distinguishing gliomas from meningiomas (*A2 vs. AGG vs. MM*) with a BER of 0.105 (p = 0.0573). R-NMU showed advantages in tasks where recursive underapproximation helped improve spectral decomposition, while G-NMU achieved perfect classification (AUC = 1.000) in certain cases but lacked statistical superiority over C-NMF. These findings suggest that while C-NMF remains the most reliable method for broad classification, NMU variants offer valuable alternatives in cases where spectral complexity challenges conventional approaches.

## Data Availability

Data can be shared within reasonable request to the corresponding author.

## Author Contributions

Conceptualization A.V. and M.J.S..; methodology A.V., M.J.S. and G.U; software, G.U; formal analysis, G.U., A.V. and M.J.S.; data curation, M.J-S.; writing—original draft preparation, M.J-S., A.V. and G.U.. All authors have read and agreed to the published version of the manuscript.

## References

1. Tate AR, Underwood J, Acosta DM, et al. Development of a decision support system for diagnosis and grading of brain tumours using in vivo magnetic resonance single voxel spectra. NMR Biomed. 2006;19(4):411–434. doi:10.1002/nbm.1016

2. Julià-Sapé M, Griffiths JR, Tate AR, et al. Classification of brain tumours from MR spectra: the INTERPRET collaboration and its outcomes. NMR Biomed. 2015;28(12):1772–1787. doi:10.1002/nbm.3439

3. Julià-Sapé M, Coronel I, Majós C, et al. Prospective diagnostic performance evaluation of single-voxel 1H MRS for typing and grading of brain tumours. NMR Biomed. 2012;25(4):661–673. doi:10.1002/nbm.1782

4. Vellido A, Romero E, Julià-Sapé M, et al. Robust discrimination of glioblastomas from metastatic brain tumors on the basis of single-voxel (1)H MRS. NMR Biomed. 2012;25(6):819–828. doi:10.1002/nbm.1797

5. Tate AR, Griffiths JR, Martínez-Pérez I, et al. Towards a method for automated classification of 1H MRS spectra from brain tumours. NMR Biomed. 1998;11(4-5):177–191. doi:10.1002/(sici)1099-1492(199806/08)11:4/5<177::aid-nbm534>3.0.co;2-u

6. Hyvarinen A. Fast and robust fixed-point algorithms for independent component analysis. IEEE Trans Neural Netw. 1999;10(3):626–634. doi:10.1109/72.761722

7. Ladroue C, Howe FA, Griffiths JR, Tate AR. Independent component analysis for automated decomposition of in vivo magnetic resonance spectra. Magn Reson Med. 2003;50(4):697–703. doi:10.1002/mrm.10595

8. Yang G, Raschke F, Barrick TR, Howe FA. Manifold Learning in MR spectroscopy using nonlinear dimensionality reduction and unsupervised clustering. Magn Reson Med. 2015;74(3):868–878. doi:10.1002/mrm.25447

9. Hao J, Zou X, Wilson M, et al. A hybrid method of application of independent component analysis to in vivo 1H MR spectra of childhood brain tumours. NMR Biomed. 2012;25(4):594–606. doi:10.1002/nbm.1776

10. Menze BH, Lichy MP, Bachert P, Kelm BM, Schlemmer HP, Hamprecht FA. Optimal classification of long echo time in vivo magnetic resonance spectra in the detection of recurrent brain tumors. NMR Biomed. 2006;19(5):599–609. doi:10.1002/nbm.1041

11. Goryawala MZ, Sheriff S, Stoyanova R, Maudsley AA. Spectral decomposition for resolving partial volume effects in MRSI. Magn Reson Med. 2018;79(6):2886–2895. doi:10.1002/mrm.26991

12. Stoyanova R, Kuesel AC, Brown TR. Application of Principal-Component Analysis for NMR Spectral Quantitation. J Magn Reson A. 1995;115(2):265–269. doi:10.1006/jmra.1995.1177

13. Kalyanam R, Boutte D, Gasparovic C, Hutchison KE, Calhoun VD. Group independent component analysis of MR spectra. Brain Behav. 2013;3(3):229–242. doi:10.1002/brb3.131

14. Szabo de Edelenyi F, Simonetti AW, Postma G, Huo R, Buydens LMC. Application of independent component analysis to 1H MR spectroscopic imaging exams of brain tumours. Anal Chim Acta. 2005;544(1):36–46. doi:10.1016/j.aca.2005.04.007

15. Stamatelatou A, Bertinetto CG, Jansen JJ, et al. A multivariate curve resolution analysis of multicenter proton spectroscopic imaging of the prostate for cancer localization and assessment of aggressiveness. NMR Biomed. 2024;37(3):e5062. doi:10.1002/nbm.5062

16. Lee DD, Seung HS. Learning the parts of objects by non-negative matrix factorization. Nature. 1999;401(6755):788-791. doi:10.1038/44565

17. Paatero P, Tapper U. Positive matrix factorization: A non-negative factor model with optimal utilization of error estimates of data values. Environmetrics. 1994;5(2):111–126. doi:10.1002/env.3170050203

18. Ding CHQ, Li T, Jordan MI. Convex and Semi-Nonnegative Matrix Factorizations. IEEE Trans Pattern Anal Mach Intell. 2010;32(1):45–55. doi:10.1109/TPAMI.2008.277

19. Vilamala A, Lisboa PJG, Ortega-Martorell S, Vellido A. Discriminant Convex Non-negative Matrix Factorization for the classification of human brain tumours. Pattern Recognit Lett. 2013;34(14):1734–1747. doi:10.1016/j.patrec.2013.05.023

20. Sajda P, Du S, Brown TR, et al. Nonnegative matrix factorization for rapid recovery of constituent spectra in magnetic resonance chemical shift imaging of the brain. IEEE Trans Med Imaging. 2004;23(12):1453–1465. doi:10.1109/TMI.2004.834626

21. Ortega-Martorell S, Lisboa PJ, Vellido A, Julià-Sapé M, Arús C. Non-negative matrix factorisation methods for the spectral decomposition of MRS data from human brain tumours. BMC Bioinformatics. 2012;13(1):38. doi:10.1186/1471-2105-13-38

22. Shungu DC, Du S, Mao X, Heier LA, Pannullo SC, Sajda P. Automated analysis of 1H magnetic resonance metabolic imaging data as an aid to clinical decision-making in the evaluation of intracranial lesions. Annu Int Conf IEEE Eng Med Biol Soc IEEE Eng Med Biol Soc Annu Int Conf. 2007;2007:4327–4330. doi:10.1109/IEMBS.2007.4353294

23. Ungan G, Pons-Escoda A, Ulinic D, et al. Early pseudoprogression and progression lesions in glioblastoma patients are both metabolically heterogeneous. NMR Biomed. 2024;37(4):e5095. doi:10.1002/nbm.5095

24. Gillis N, Glineur F. Using underapproximations for sparse nonnegative matrix factorization. Pattern Recognit. 2010;43(4):1676–1687. doi:10.1016/j.patcog.2009.11.013

25. Gillis N, Plemmons RJ. Dimensionality reduction, classification, and spectral mixture analysis using nonnegative underapproximation. 2010;7695:76951A. doi:10.1117/12.849345

26. Gillis N, Plemmons RJ. Sparse nonnegative matrix underapproximation and its application to hyperspectral image analysis. Linear Algebra Its Appl. 2013;438(10):3991–4007. doi:10.1016/j.laa.2012.04.033

27. Tepper M, Sapiro G. Nonnegative Matrix Underapproximation for Robust Multiple Model Fitting. In: 2017 IEEE Conference on Computer Vision and Pattern Recognition (CVPR).; 2017:655–663. doi:10.1109/CVPR.2017.77

28. Ungan G, Arús C, Vellido A, Julià-Sapé M. A comparison of non-negative matrix underapproximation methods for the decomposition of magnetic resonance spectroscopy data from human brain tumors. NMR Biomed. Published online August 15, 2023:e5020. doi:10.1002/nbm.5020

29. Julià-Sapé M, Acosta D, Mier M, Arùs C, Watson D, INTERPRET consortium. A multi-centre, web-accessible and quality control-checked database of in vivo MR spectra of brain tumour patients. Magma N Y N. 2006;19(1):22–33. doi:10.1007/s10334-005-0023-x

30. García-Gómez JM, Luts J, Julià-Sapé M, et al. Multiproject–multicenter evaluation of automatic brain tumor classification by magnetic resonance spectroscopy. Magn Reson Mater Phys Biol Med. 2009;22(1):5–18. doi:10.1007/s10334-008-0146-y

31. Lukas L, Devos A, Suykens JAK, et al. Brain tumor classification based on long echo proton MRS signals. Artif Intell Med. 2004;31(1):73–89. doi:10.1016/j.artmed.2004.01.001

32. Luts J, Poullet JB, Garcia-Gomez JM, et al. Effect of feature extraction for brain tumor classification based on short echo time 1H MR spectra. Magn Reson Med. 2008;60(2):288–298. doi:10.1002/mrm.21626

33. Devos A, Lukas L, Suykens JAK, et al. Classification of brain tumours using short echo time 1H MR spectra. J Magn Reson. 2004;170(1):164–175. doi:10.1016/j.jmr.2004.06.010

34. Cianfoni A, Law M, Re TJ, Dubowitz DJ, Rumboldt Z, Imbesi SG. Clinical pitfalls related to short and long echo times in cerebral MR spectroscopy. J Neuroradiol J Neuroradiol. 2011;38(2):69–75. doi:10.1016/j.neurad.2010.10.001

35. Zhu H, Barker PB. MR Spectroscopy and Spectroscopic Imaging of the Brain. Methods Mol Biol Clifton NJ. 2011;711:203–226. doi:10.1007/978-1-61737-992-5_9

36. Murali-Manohar S, Borbath T, Wright AM, Soher B, Mekle R, Henning A. T2 relaxation times of macromolecules and metabolites in the human brain at 9.4 T. Magn Reson Med. 2020;84(2):542–558. doi:10.1002/mrm.28174

37. Prescot AP, Shi X, Choi C, Renshaw PerryF. In Vivo T2 Relaxation Time Measurement with Echo-Time Averaging. NMR Biomed. 2014;27(8):863–869. doi:10.1002/nbm.3115

38. Liu H, Xiang QS, Tam R, et al. Introduction to a novel T2 relaxation analysis method SAME-ECOS: Spectrum Analysis for Multiple Exponentials via Experimental Condition Oriented Simulation. Published online September 14, 2020. doi:10.48550/arXiv.2009.06761

