## Supplementary Materials 1 for "Evaluating Non-Negative Matrix Underapproximation for the Analysis of Long Echo Time Magnetic Resonance Spectroscopy Data in Human Brain Tumors"

### Slide 1
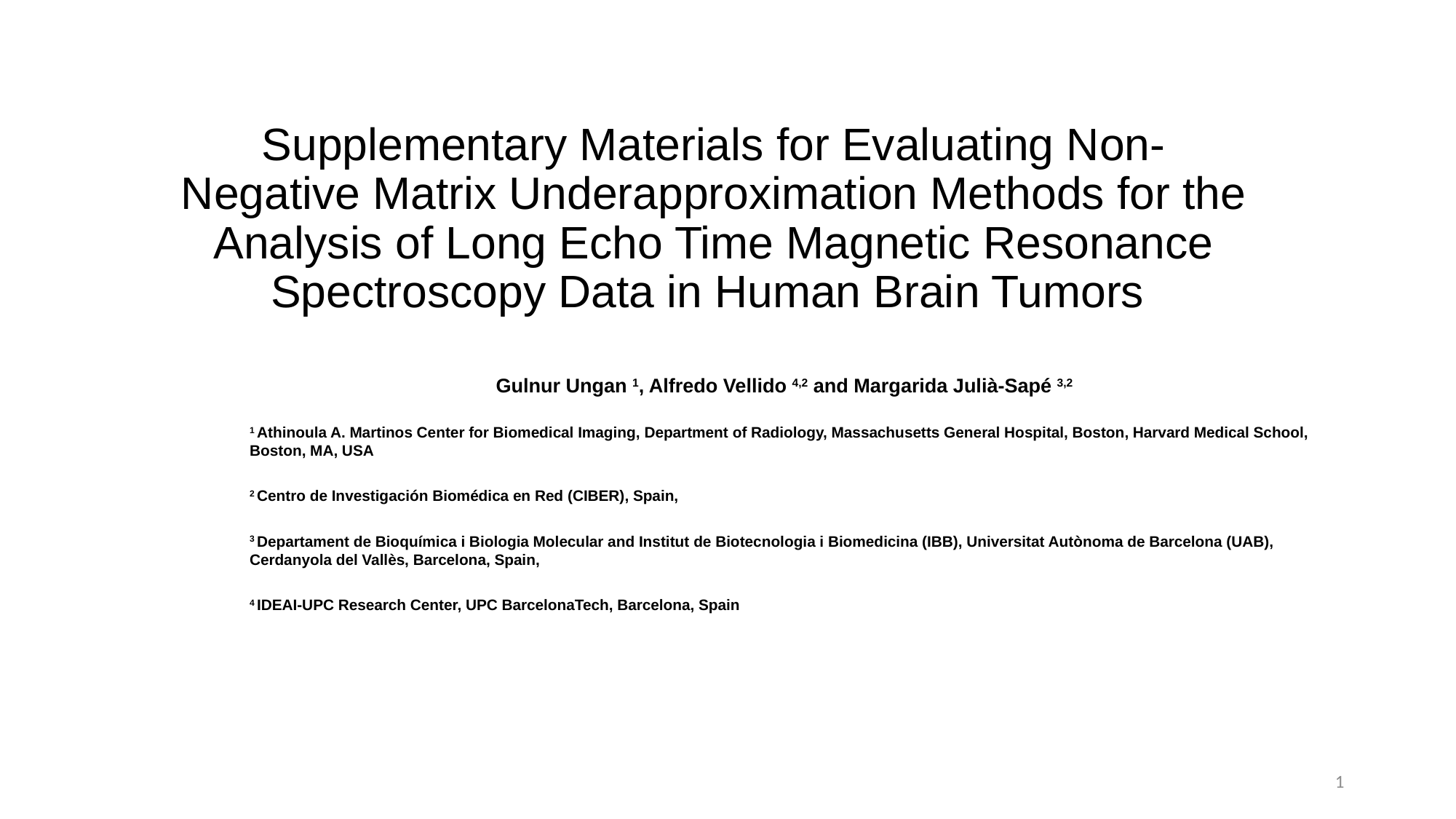

#### Supplementary Materials for Evaluating Non-Negative Matrix Underapproximation Methods for the Analysis of Long Echo Time Magnetic Resonance Spectroscopy Data in Human Brain Tumors
Gulnur Ungan ^1^, Alfredo Vellido ^4,2^ and Margarida Julià-Sapé ^3,2^
^1^ Athinoula A. Martinos Center for Biomedical Imaging, Department of Radiology, Massachusetts General Hospital, Boston, Harvard Medical School, Boston, MA, USA
^2^ Centro de Investigación Biomédica en Red (CIBER), Spain,
^3^ Departament de Bioquímica i Biologia Molecular and Institut de Biotecnologia i Biomedicina (IBB), Universitat Autònoma de Barcelona (UAB), Cerdanyola del Vallès, Barcelona, Spain,
^4^ IDEAI-UPC Research Center, UPC BarcelonaTech, Barcelona, Spain
1

### Slide 2
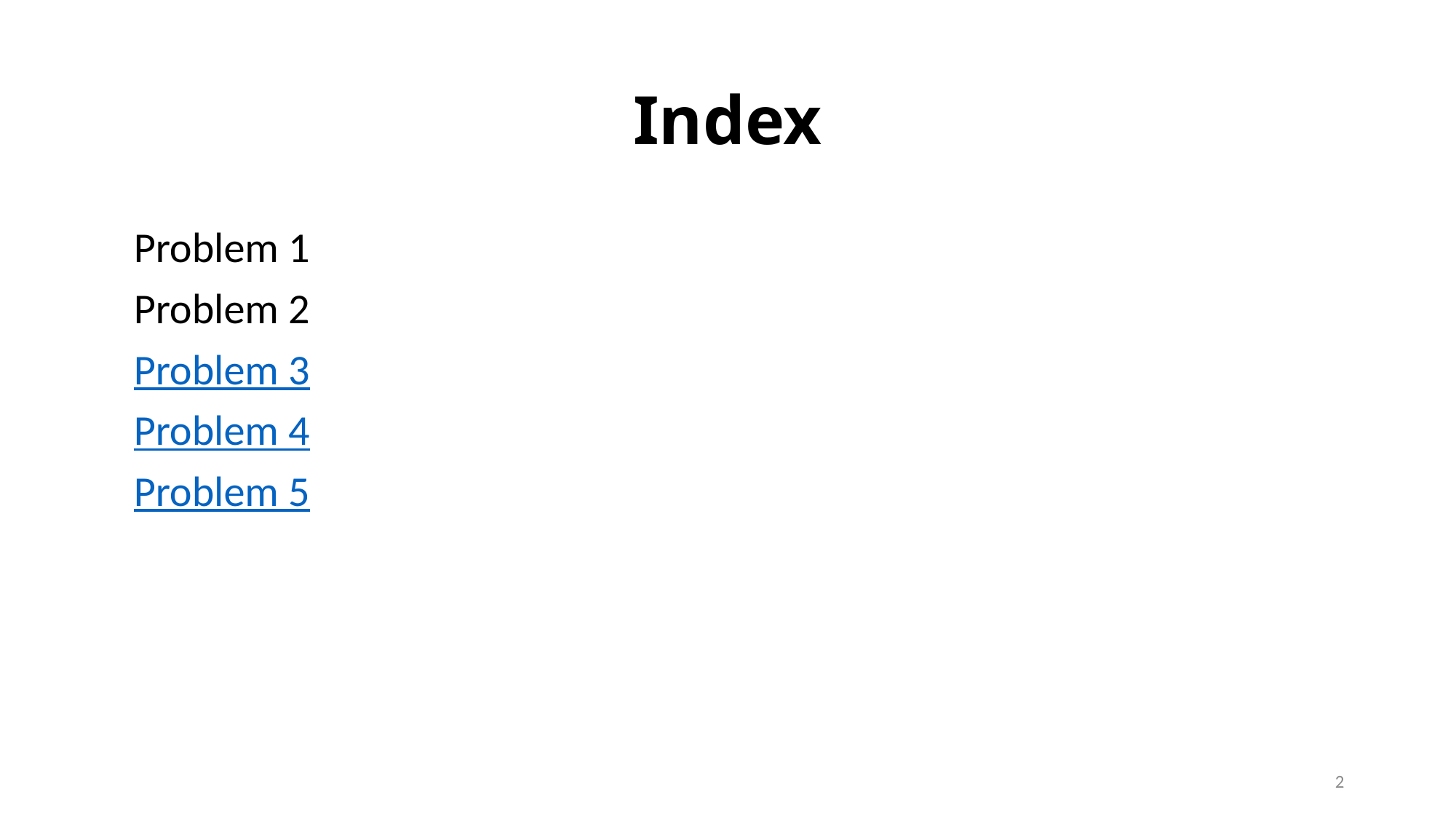

#### Index
Problem 1
Problem 2
Problem 3
Problem 4
Problem 5
2

### Slide 3
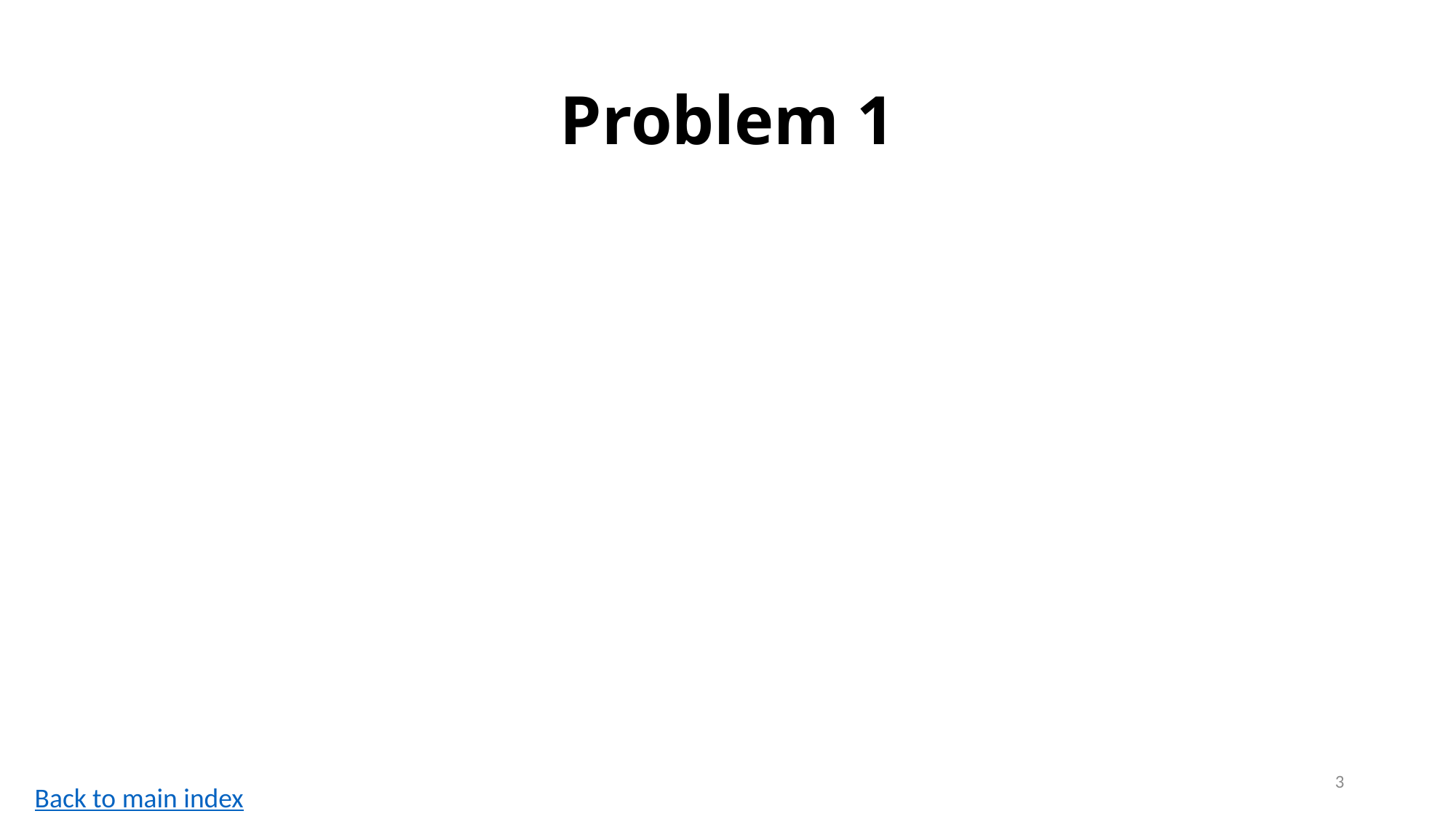

#### Problem 1
3
Back to main index

### Slide 4
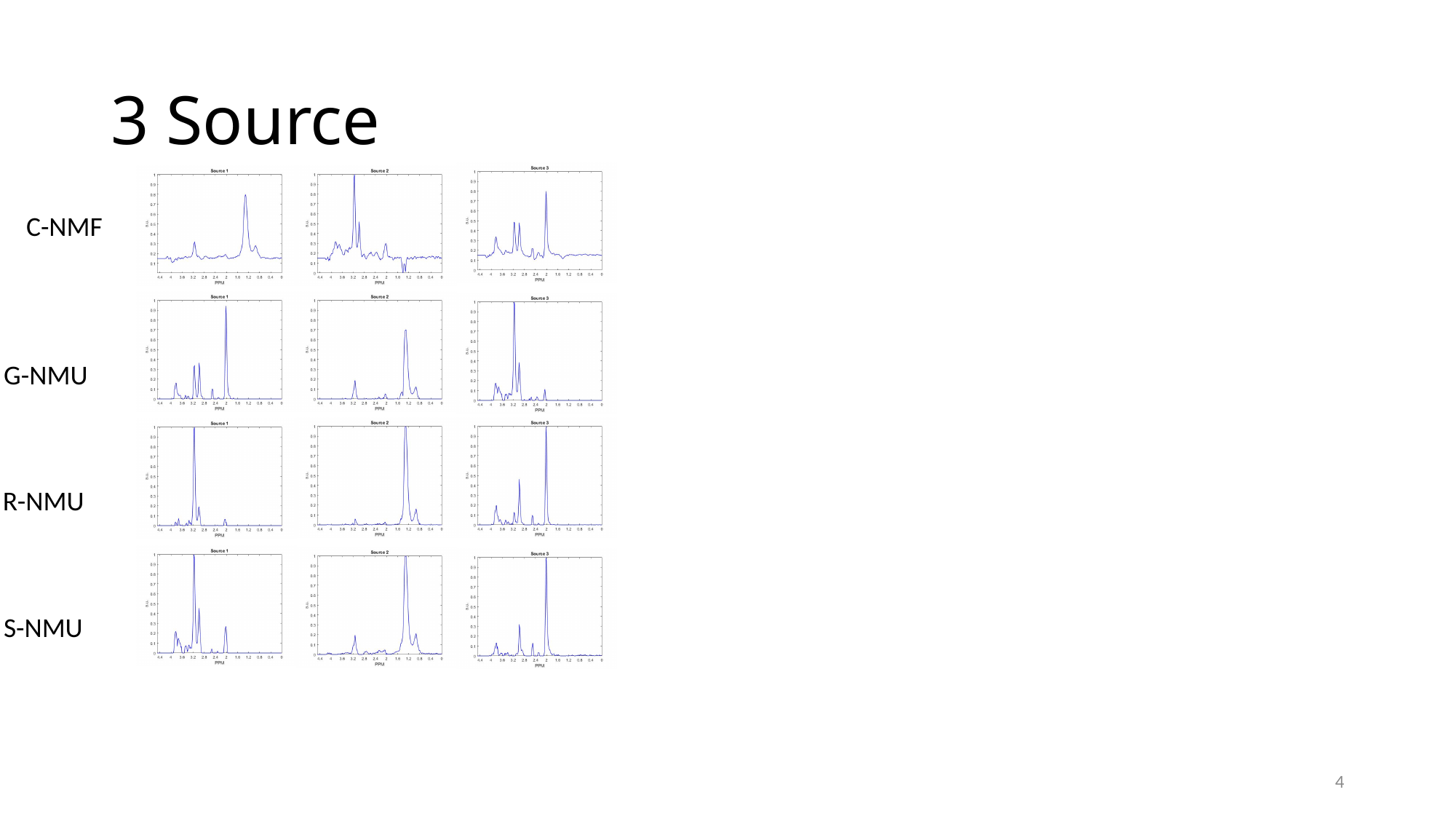

#### 3 Source
C-NMF
G-NMU
R-NMU
S-NMU
4

### Slide 5
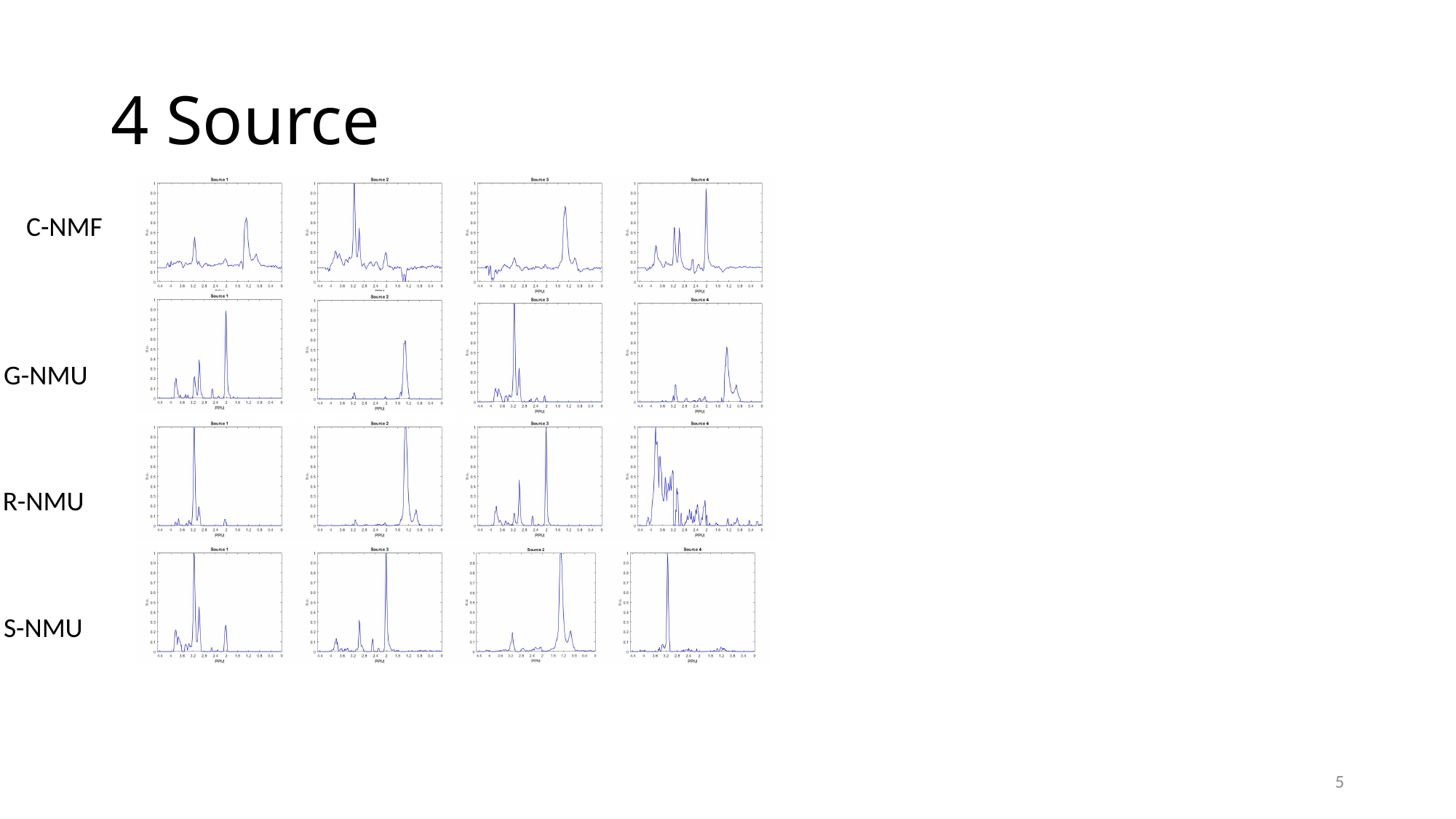

#### 4 Source
C-NMF
G-NMU
R-NMU
S-NMU
5

### Slide 6
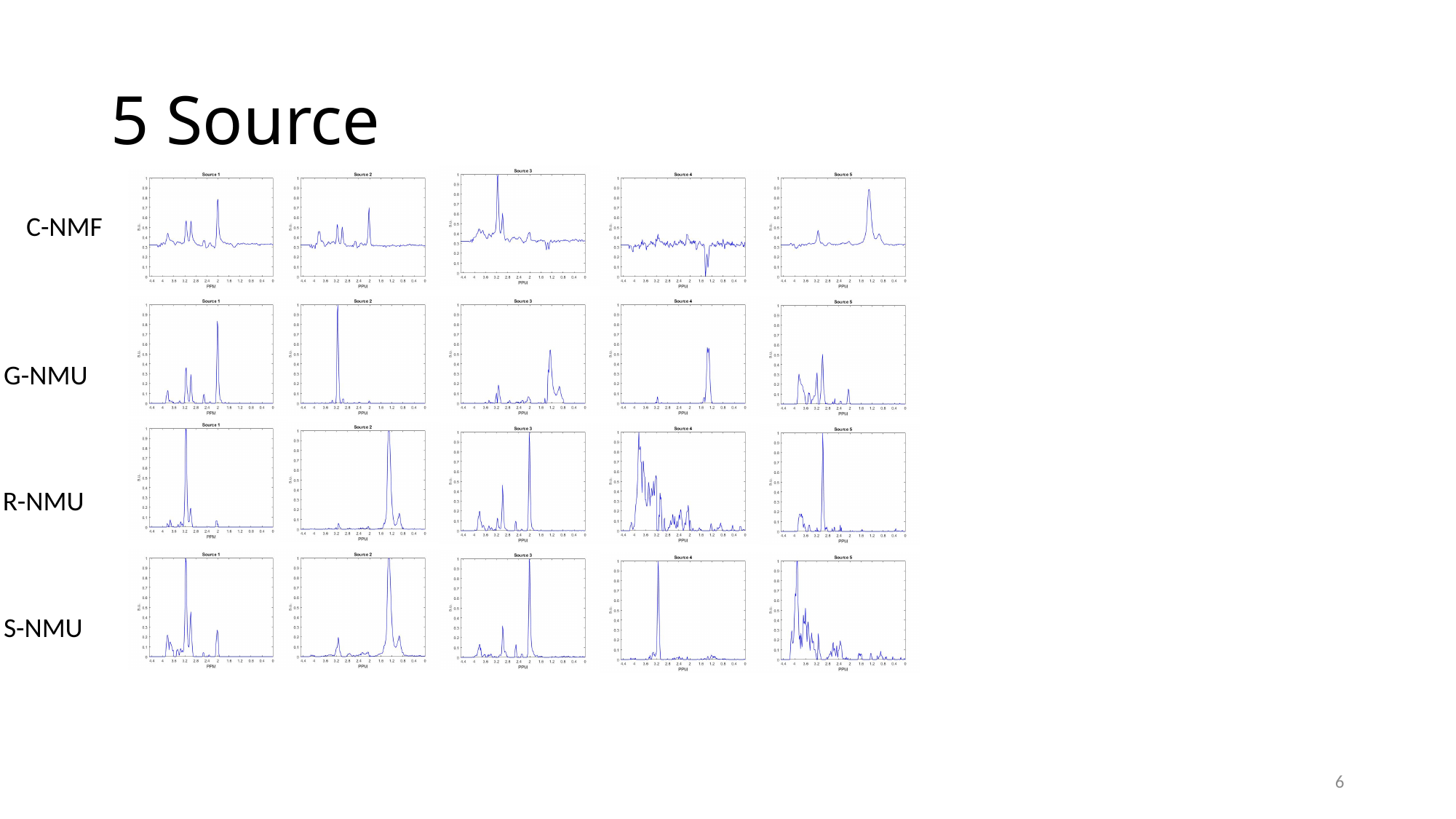

#### 5 Source
C-NMF
G-NMU
R-NMU
S-NMU
6

### Slide 7
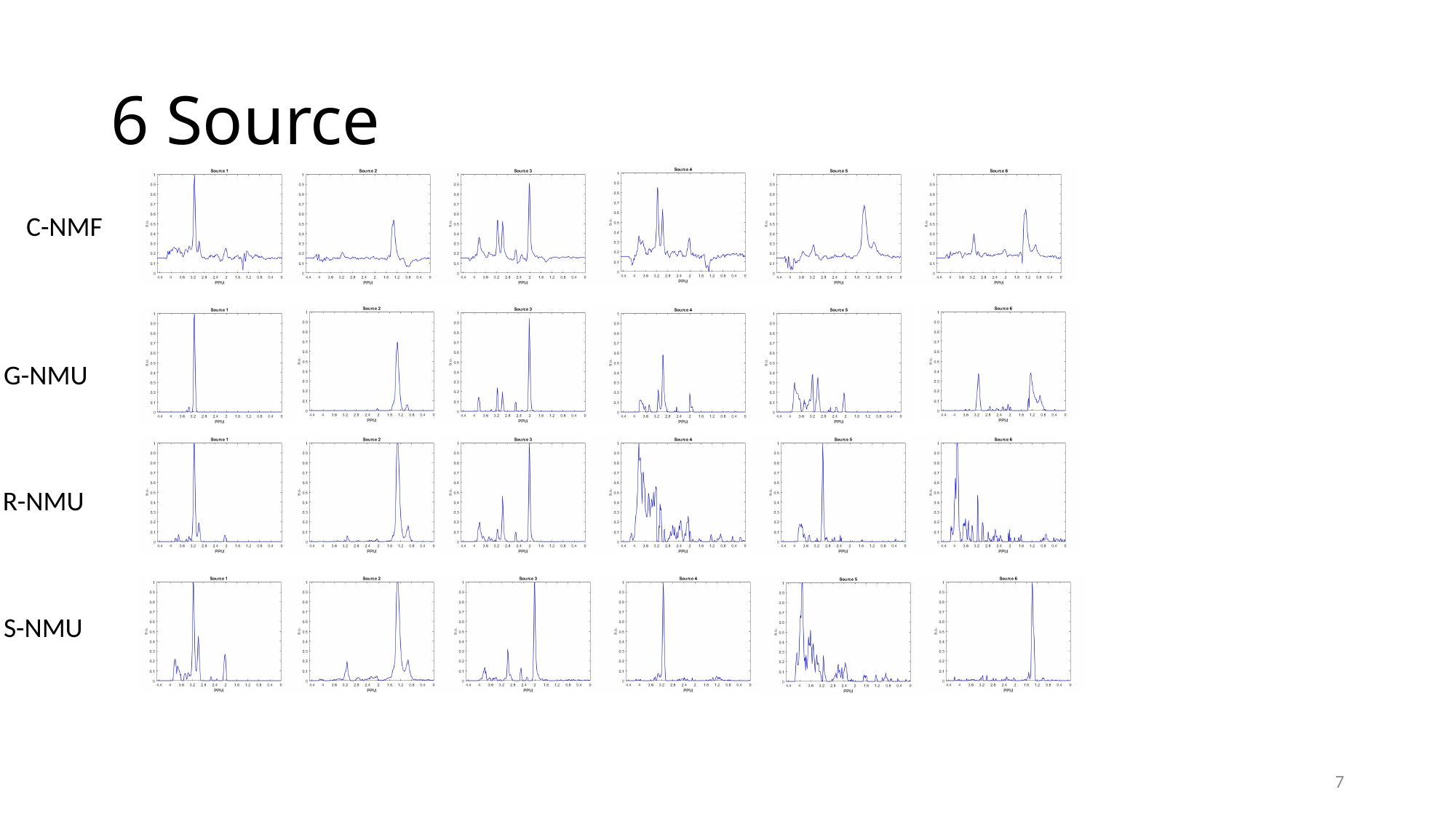

#### 6 Source
C-NMF
G-NMU
R-NMU
S-NMU
7

### Slide 8
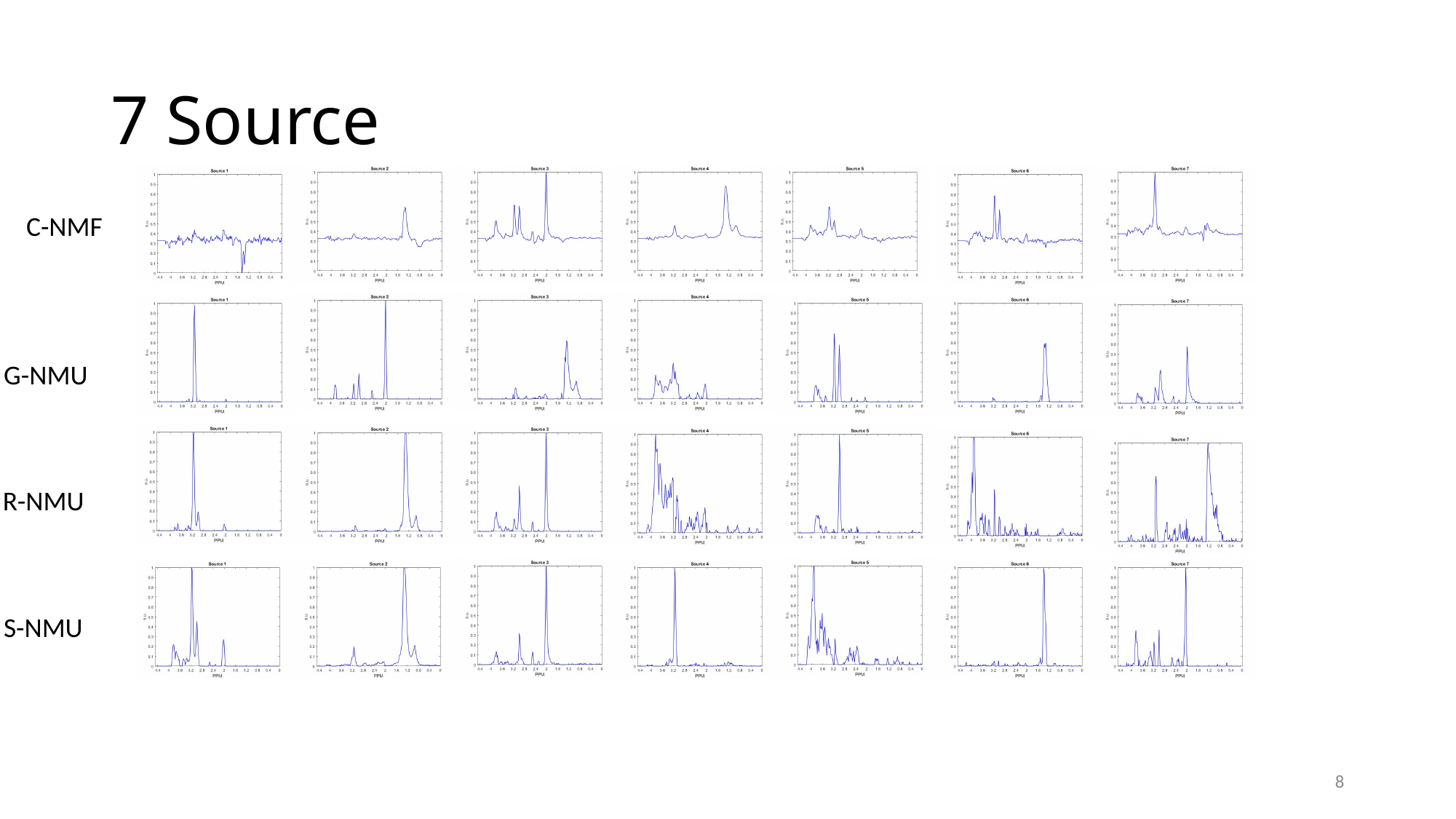

#### 7 Source
C-NMF
G-NMU
R-NMU
S-NMU
8

### Slide 9
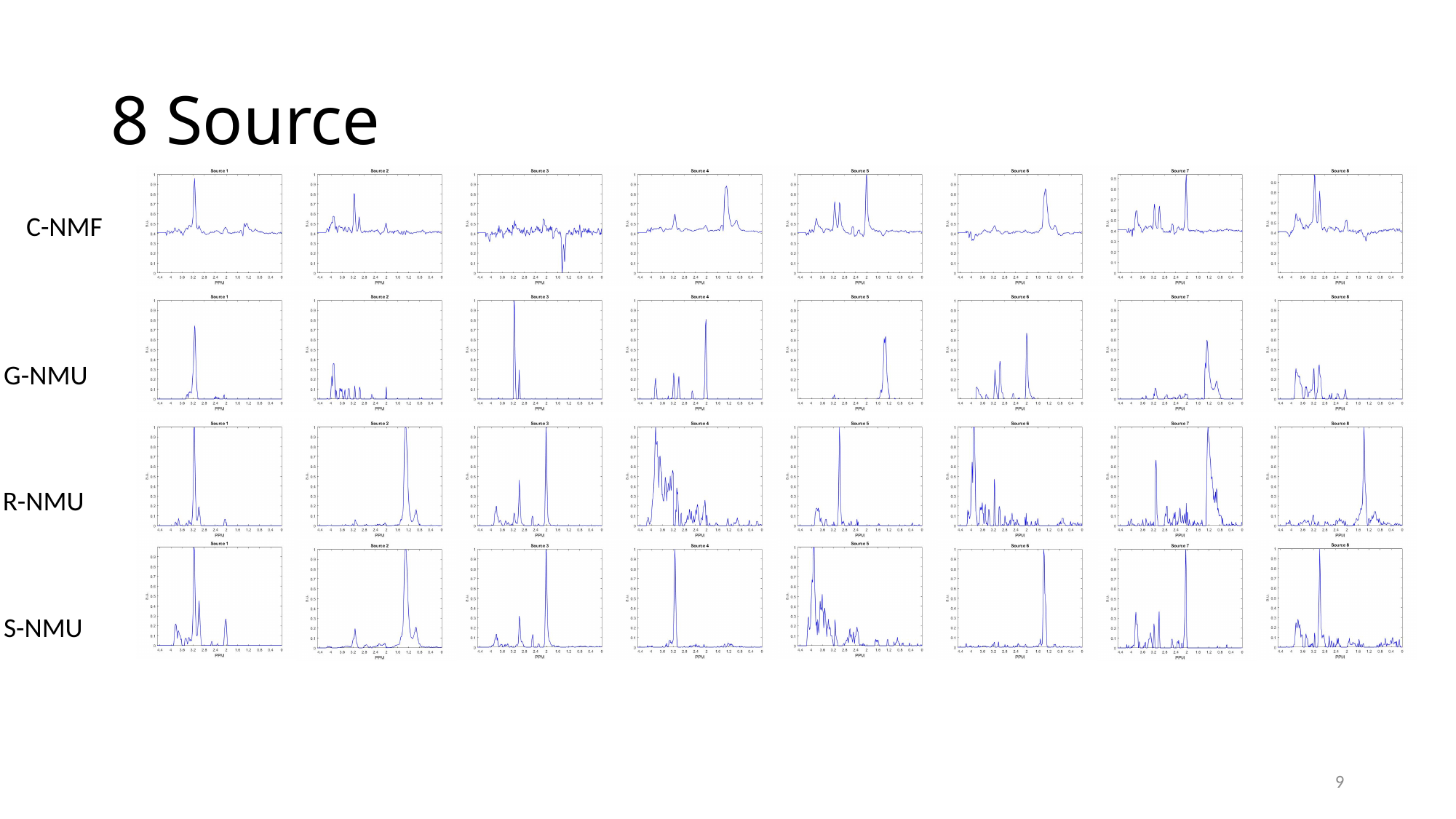

#### 8 Source
C-NMF
G-NMU
R-NMU
S-NMU
9

### Slide 10
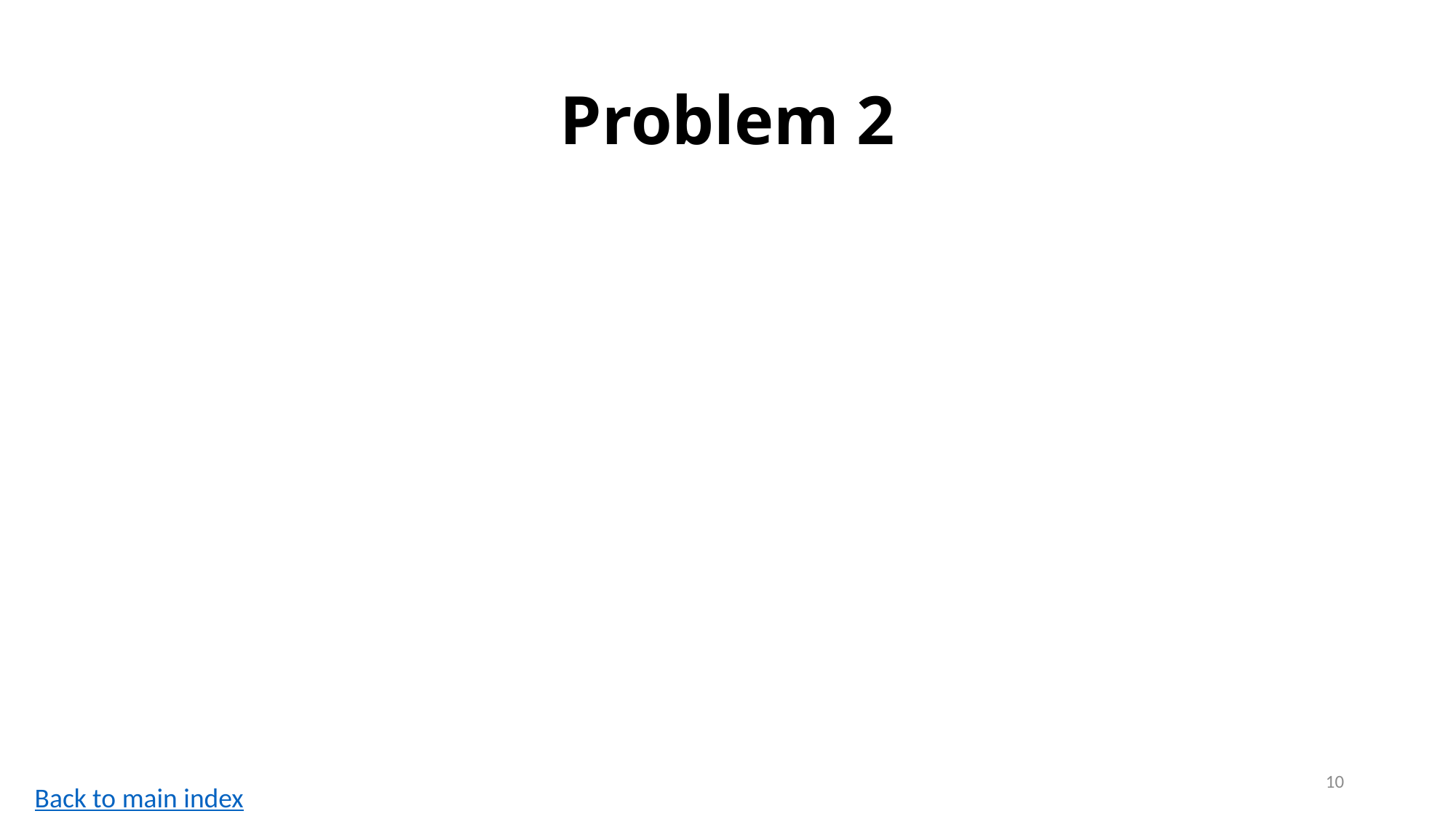

#### Problem 2
10
Back to main index

### Slide 11
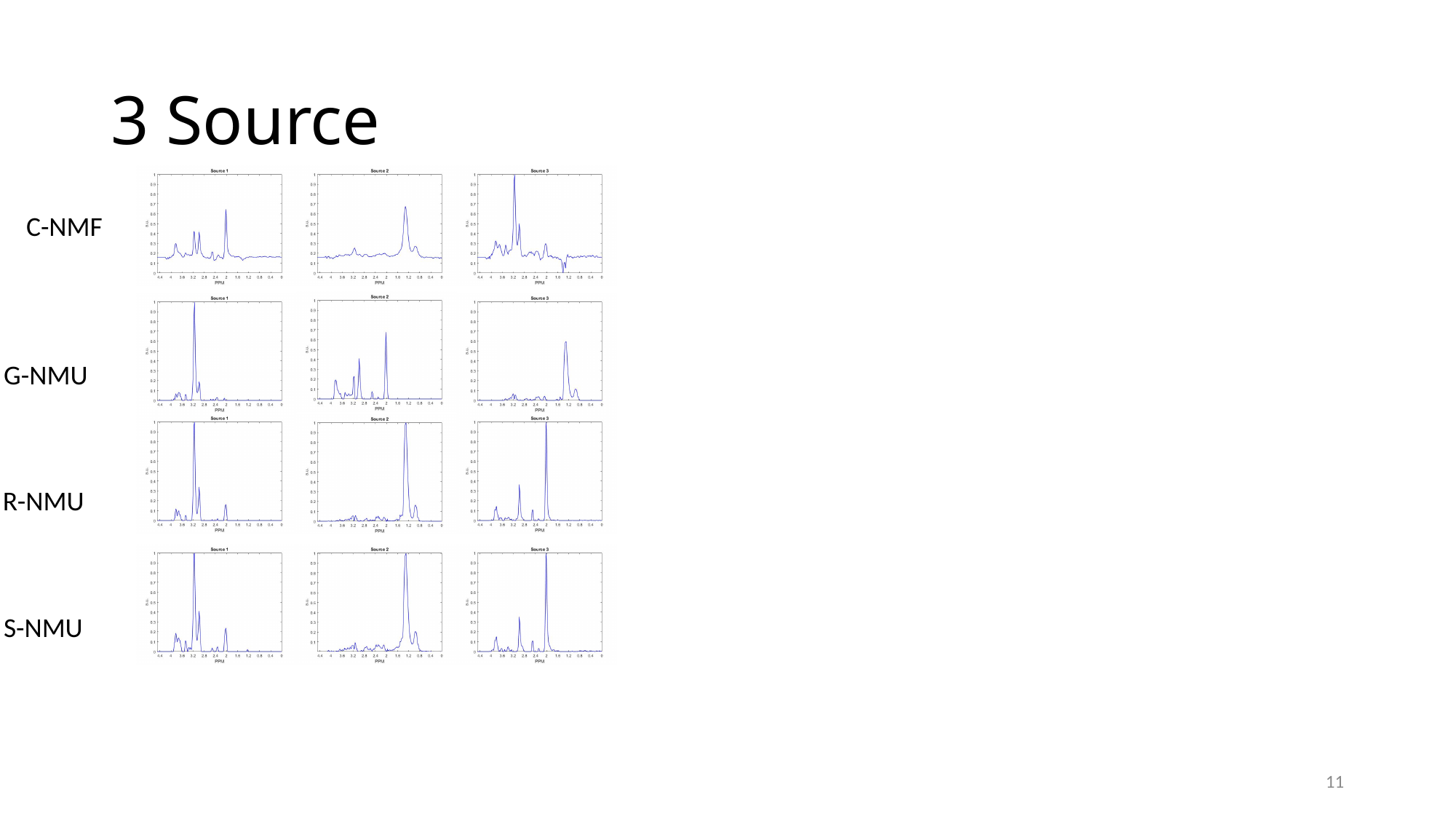

#### 3 Source
C-NMF
G-NMU
R-NMU
S-NMU
11

### Slide 12
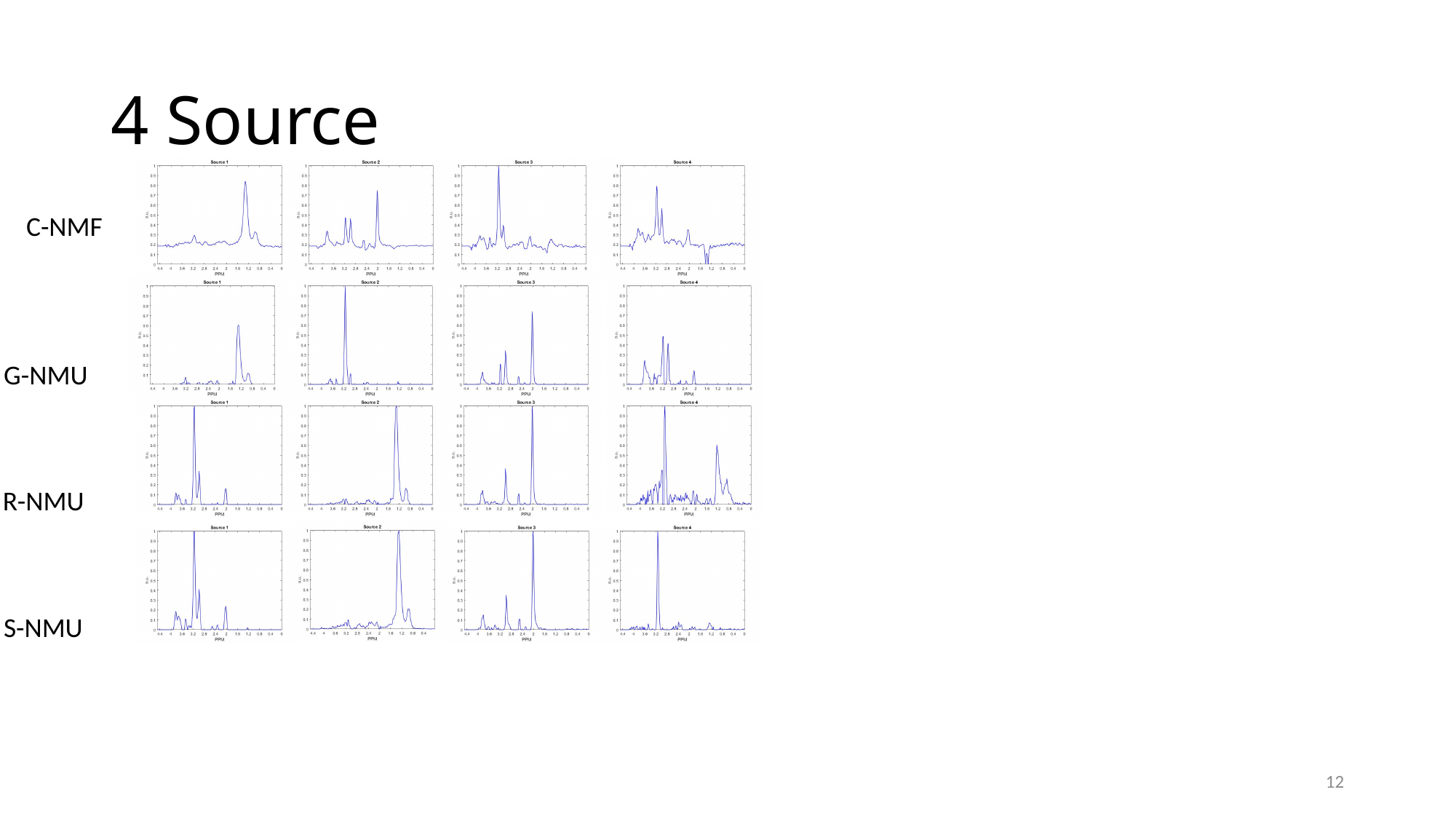

#### 4 Source
C-NMF
G-NMU
R-NMU
S-NMU
12

### Slide 13
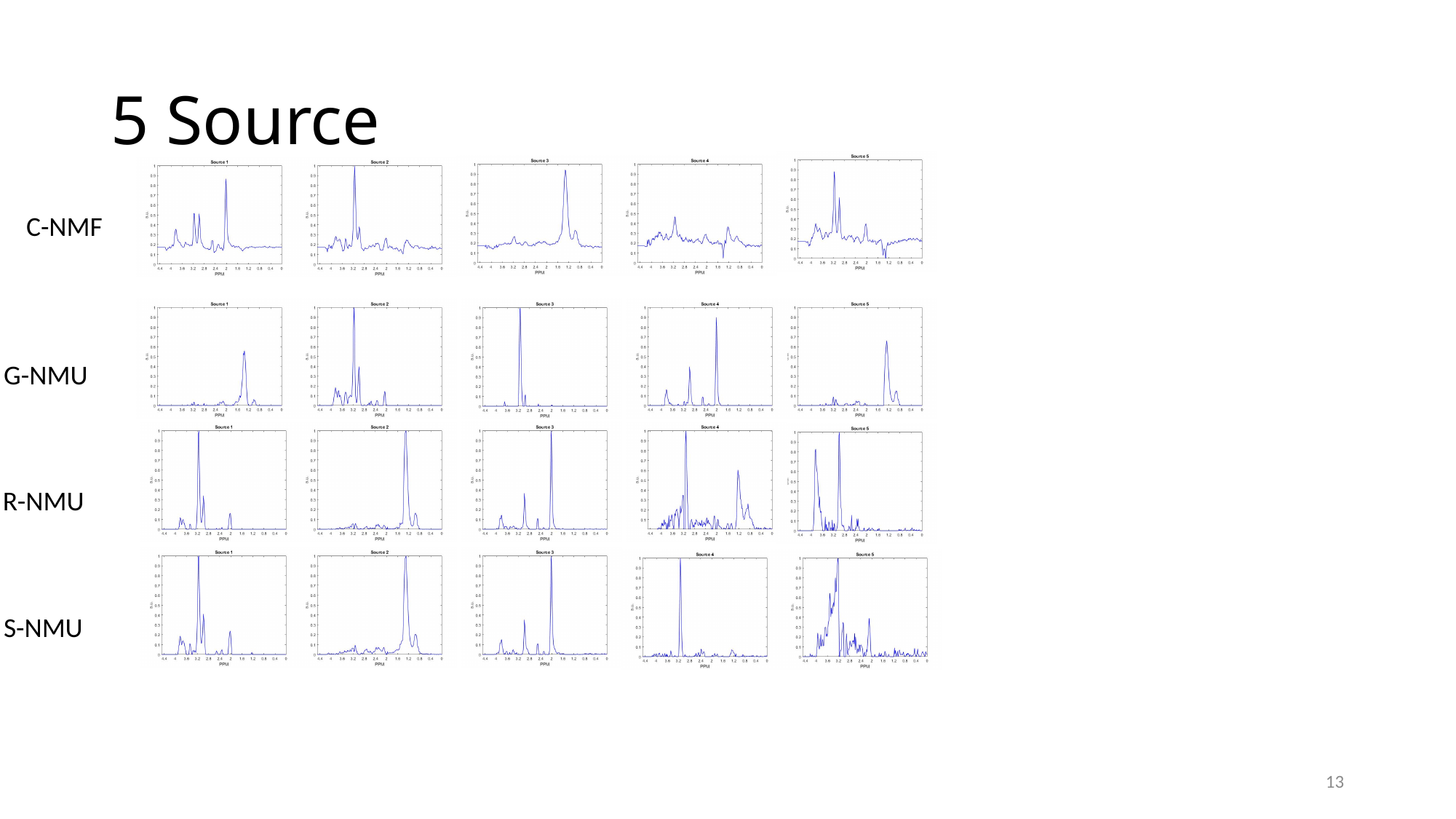

#### 5 Source
C-NMF
G-NMU
R-NMU
S-NMU
13

### Slide 14
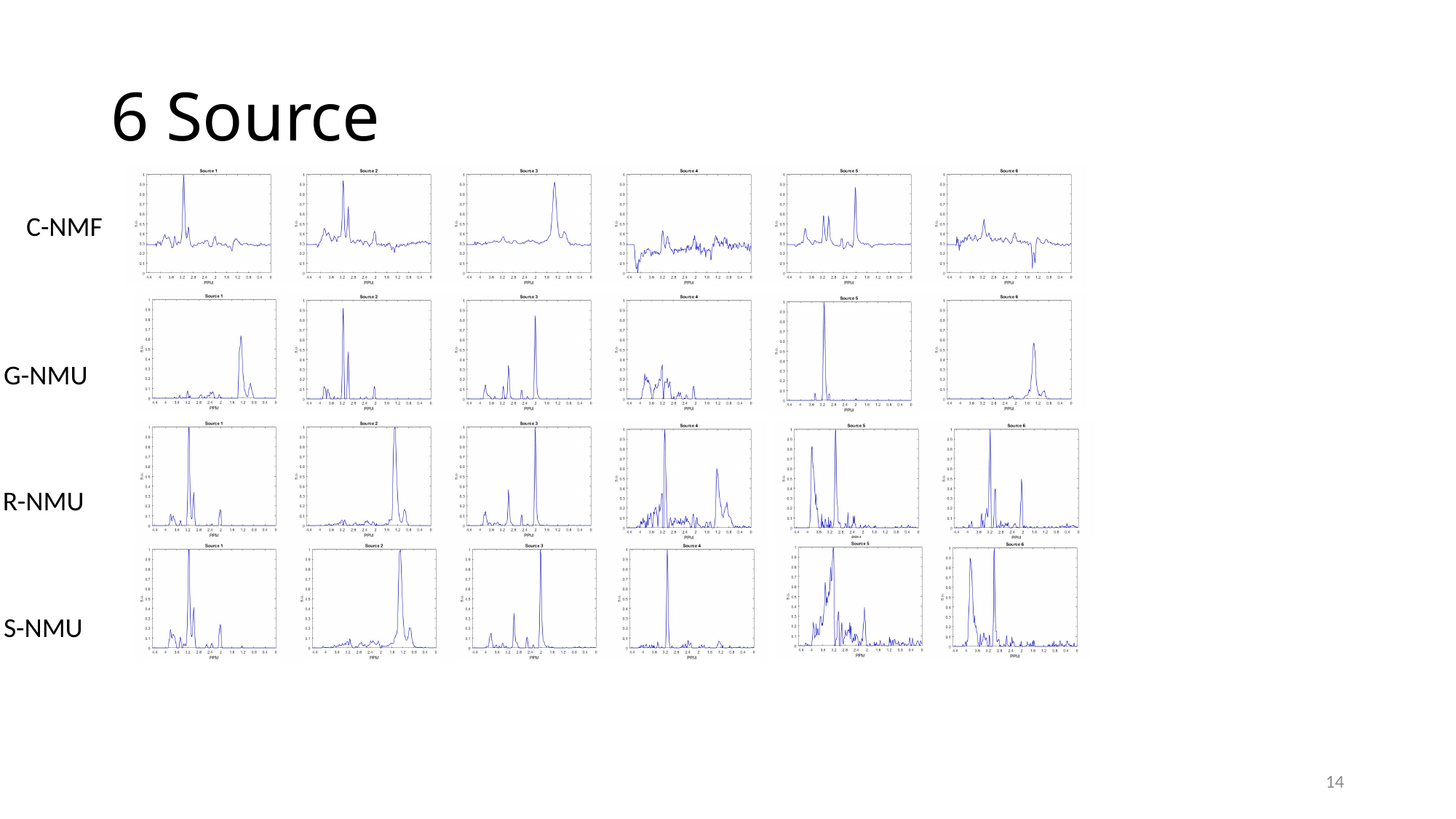

#### 6 Source
C-NMF
G-NMU
R-NMU
S-NMU
14

### Slide 15
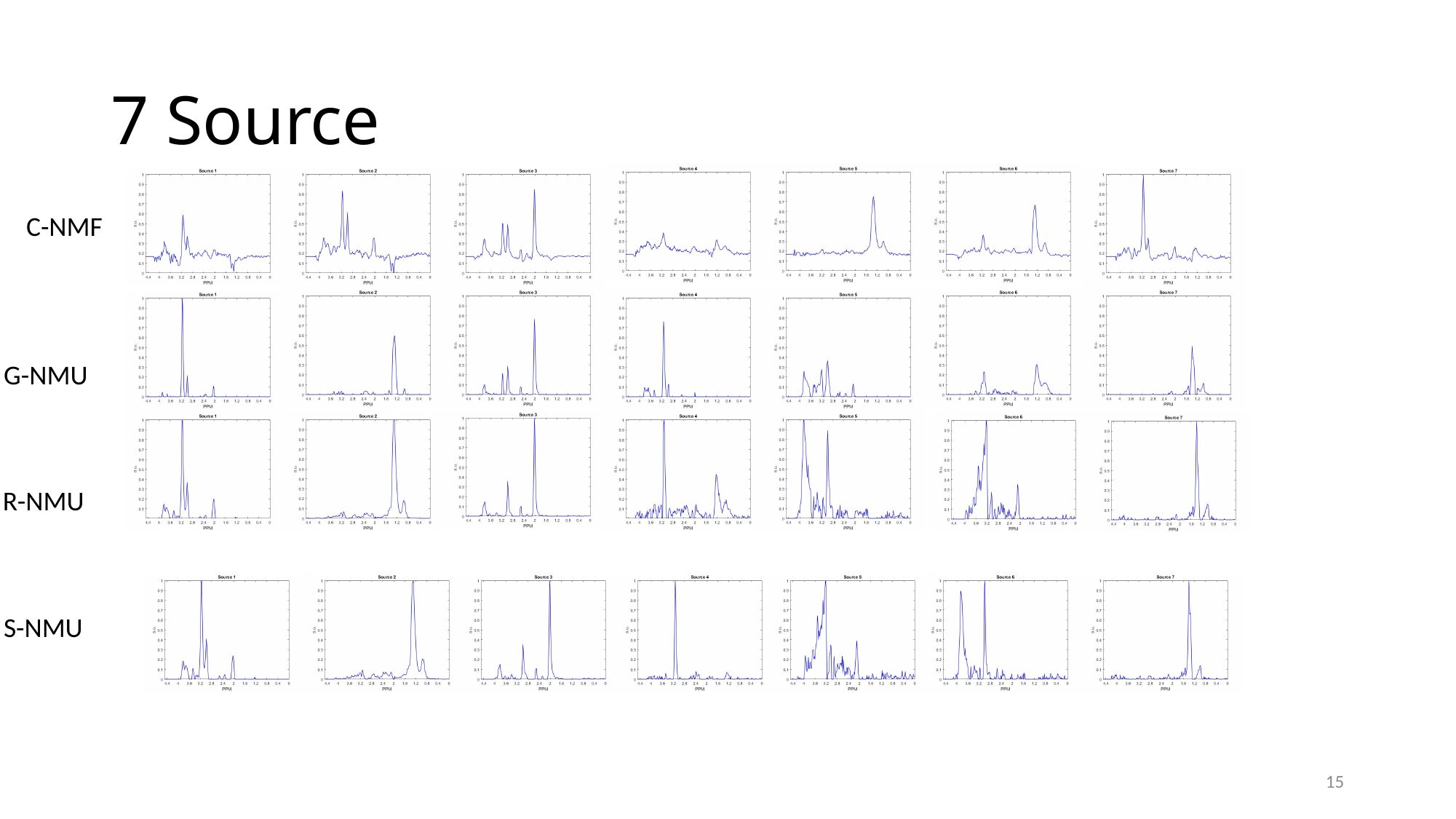

#### 7 Source
C-NMF
G-NMU
R-NMU
S-NMU
15

### Slide 16
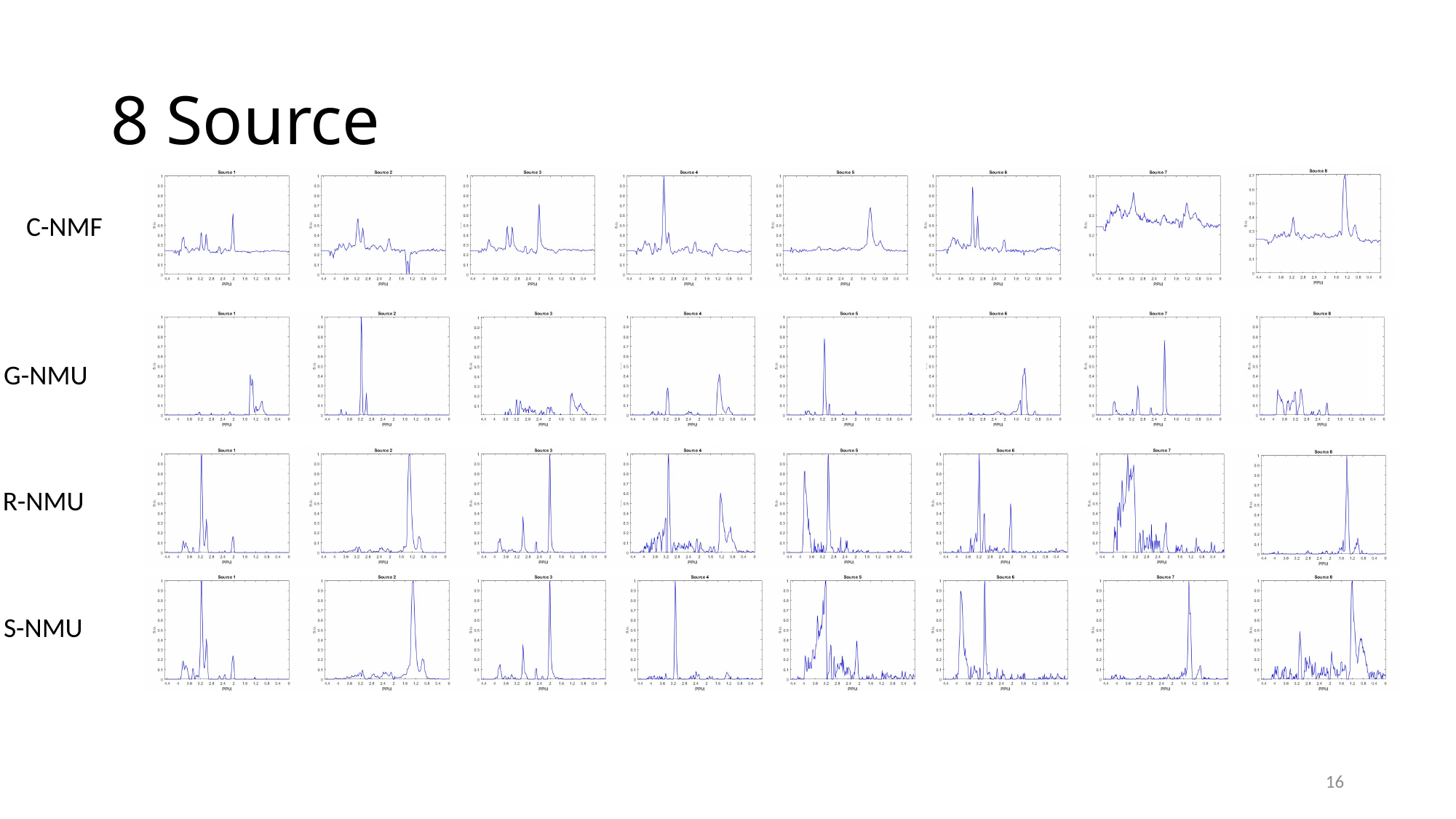

#### 8 Source
C-NMF
G-NMU
R-NMU
S-NMU
16

### Slide 17
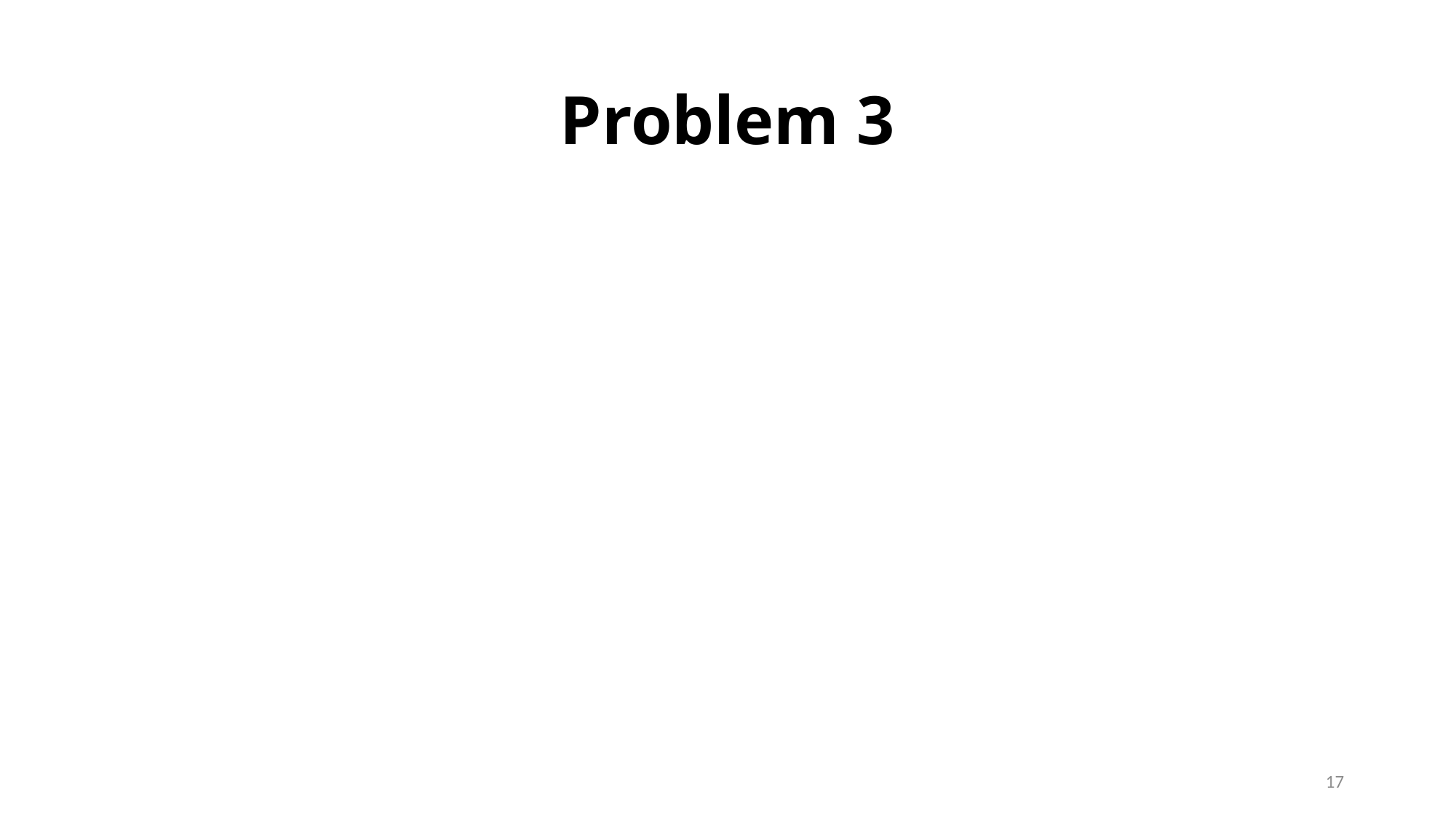

#### Problem 3
17

### Slide 18
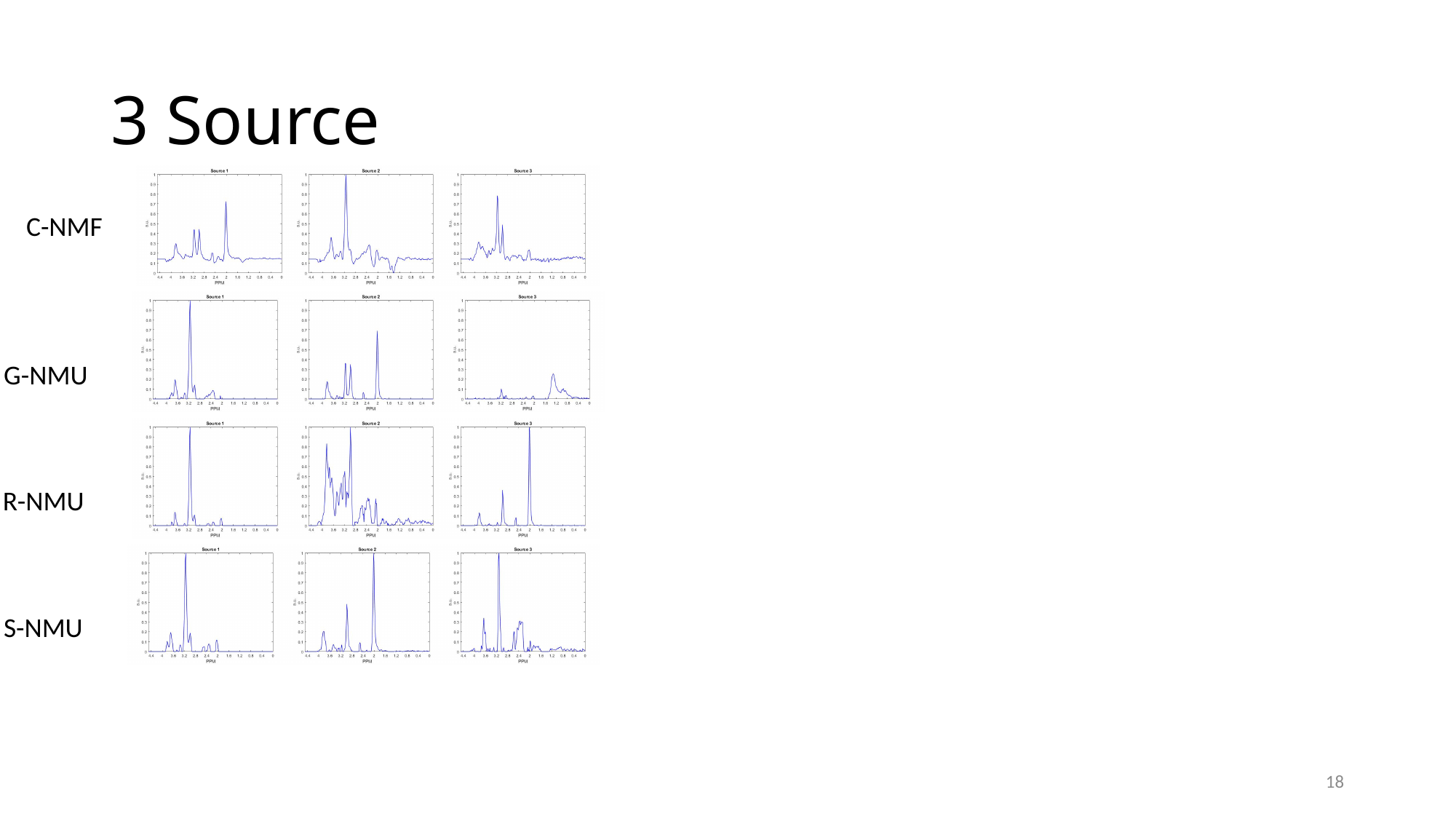

#### 3 Source
C-NMF
G-NMU
R-NMU
S-NMU
18

### Slide 19
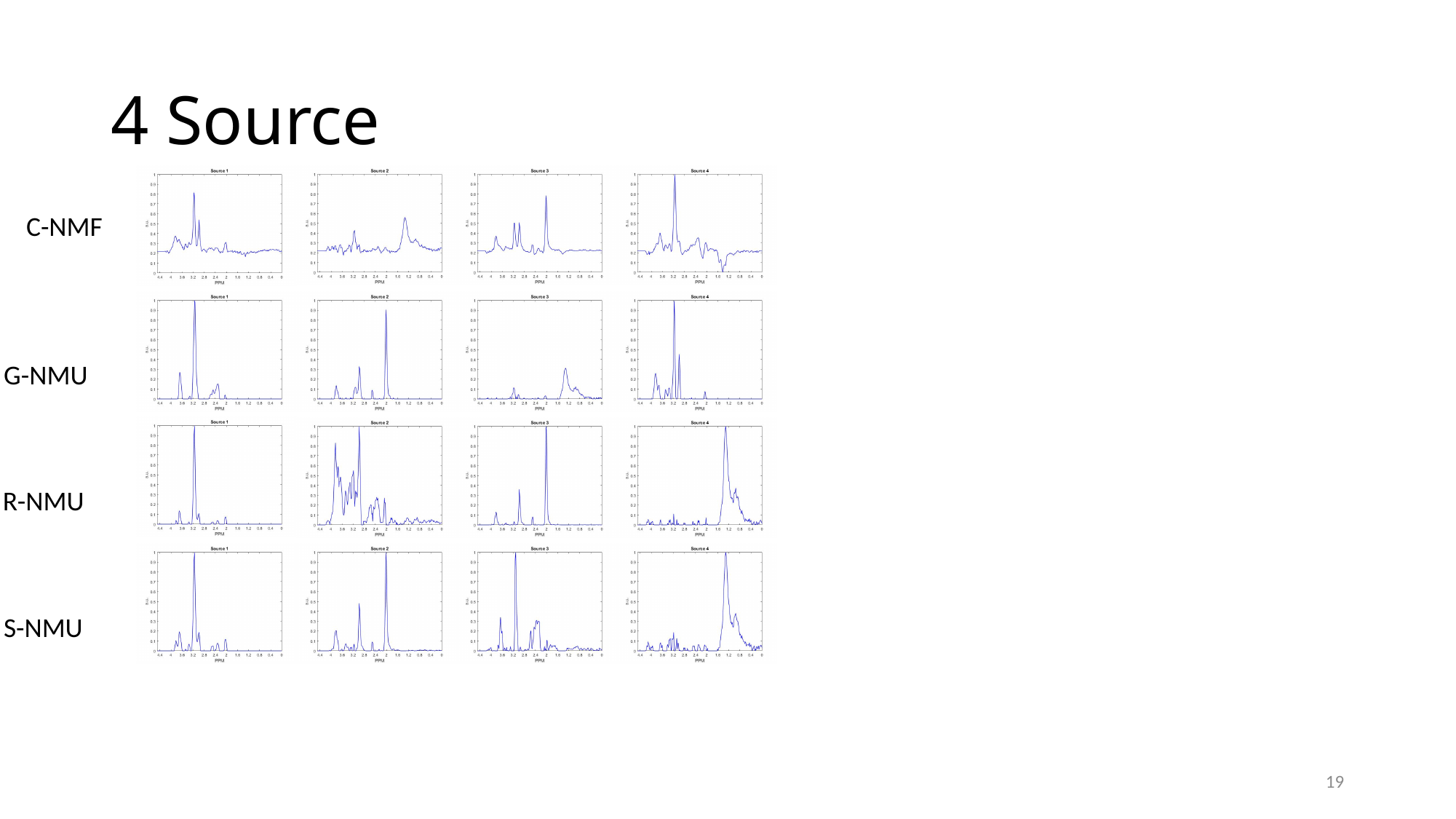

#### 4 Source
C-NMF
G-NMU
R-NMU
S-NMU
19

### Slide 20
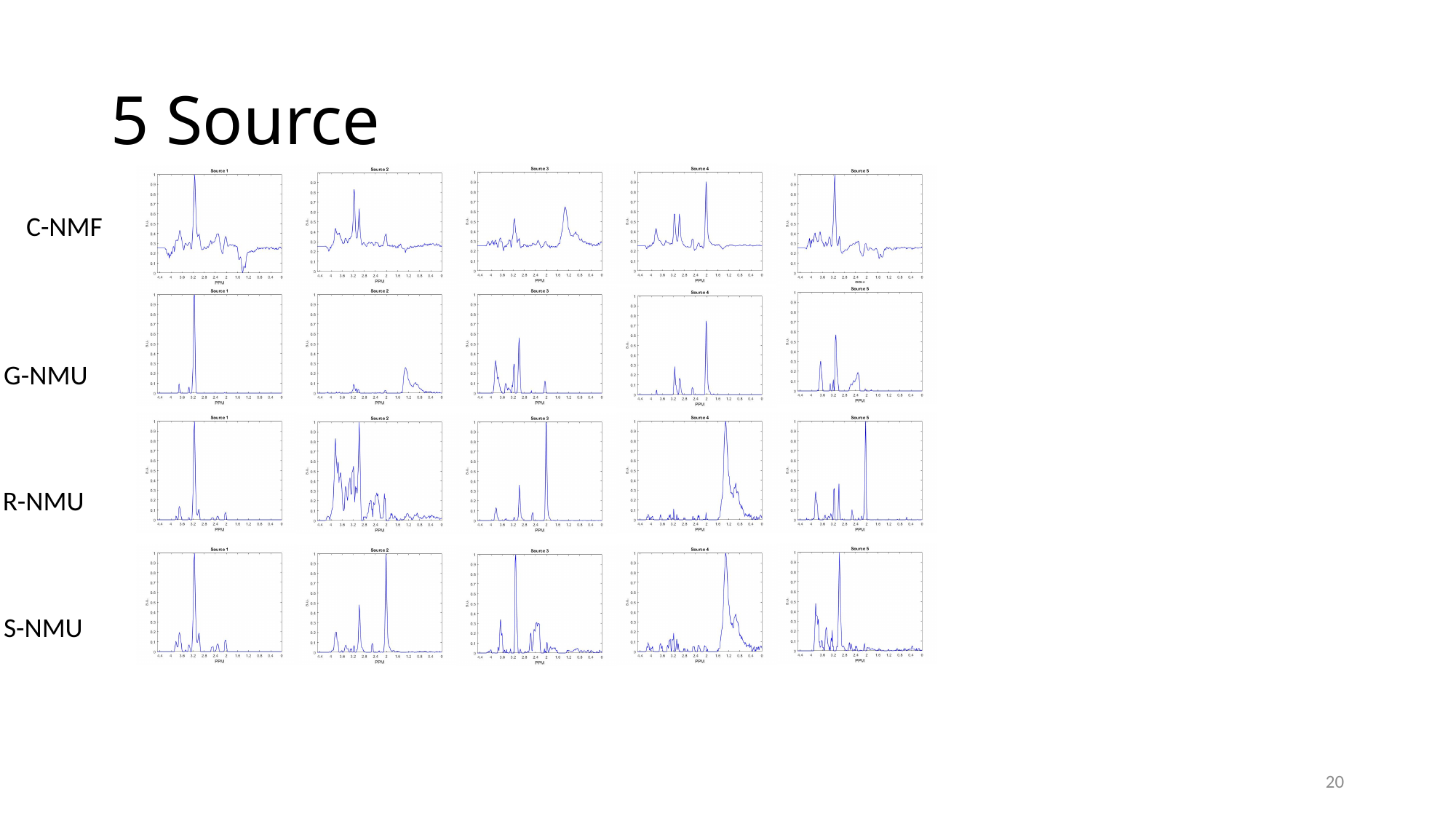

#### 5 Source
C-NMF
G-NMU
R-NMU
S-NMU
20

### Slide 21
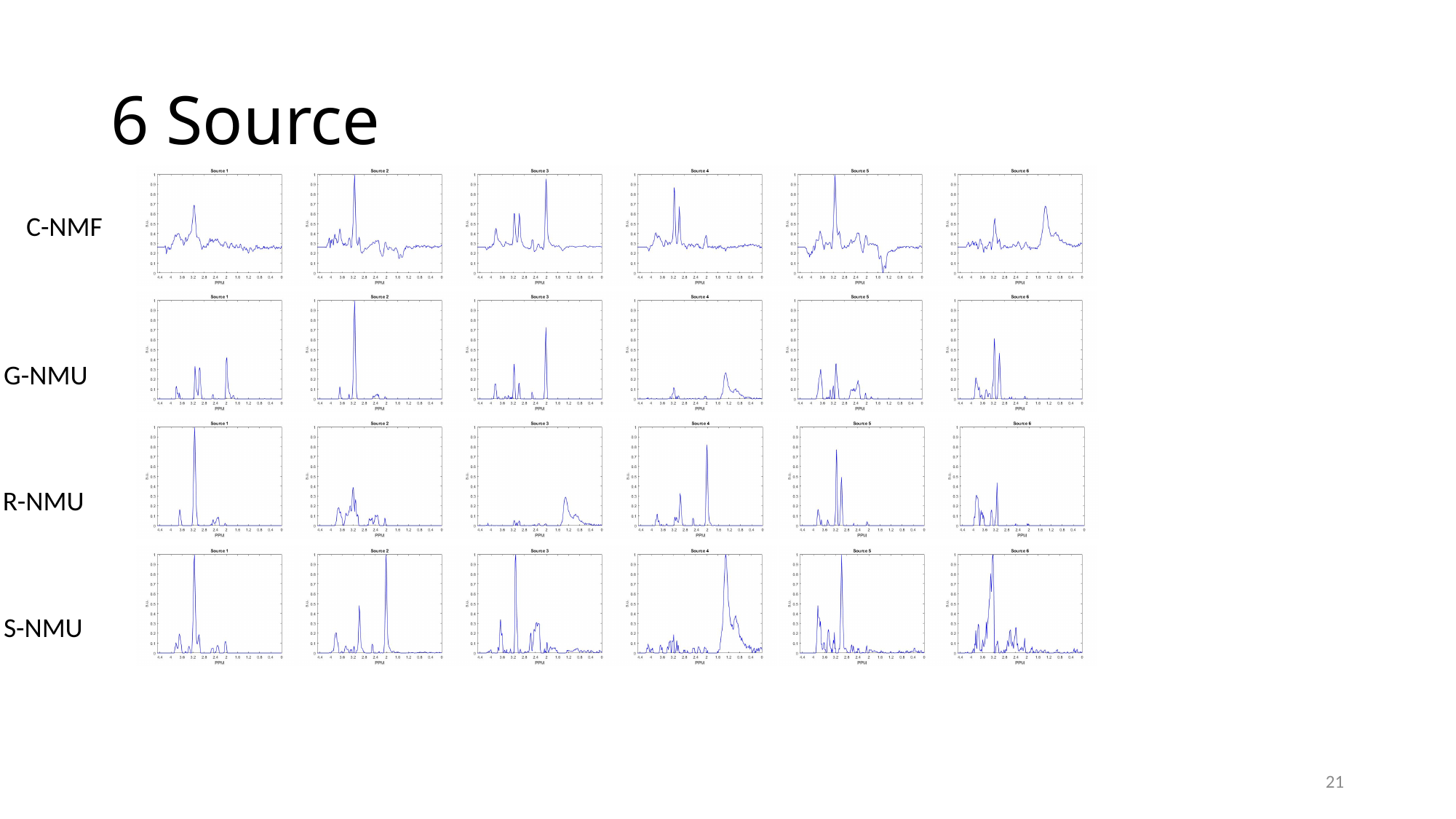

#### 6 Source
C-NMF
G-NMU
R-NMU
S-NMU
21

### Slide 22
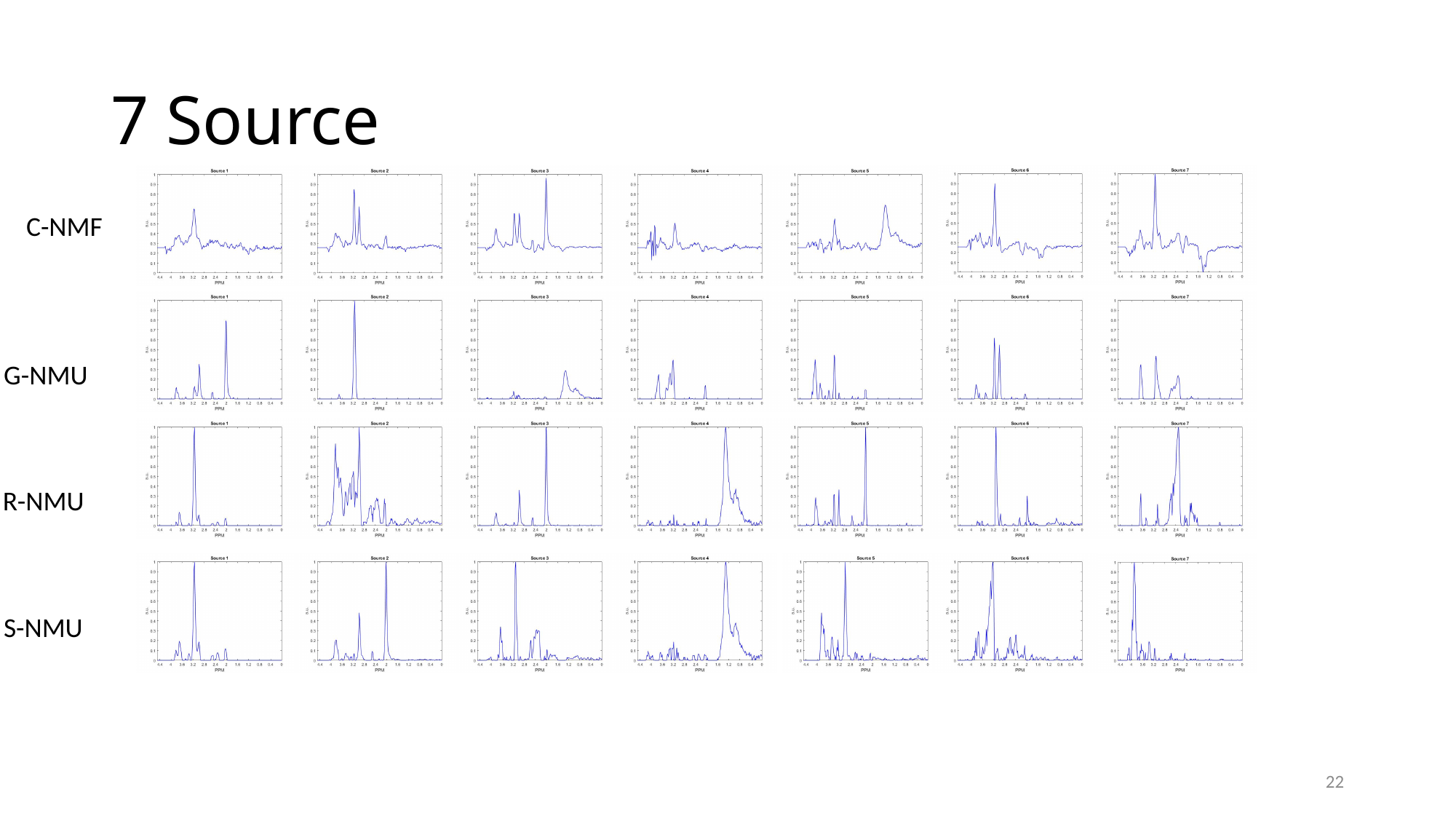

#### 7 Source
C-NMF
G-NMU
R-NMU
S-NMU
22

### Slide 23
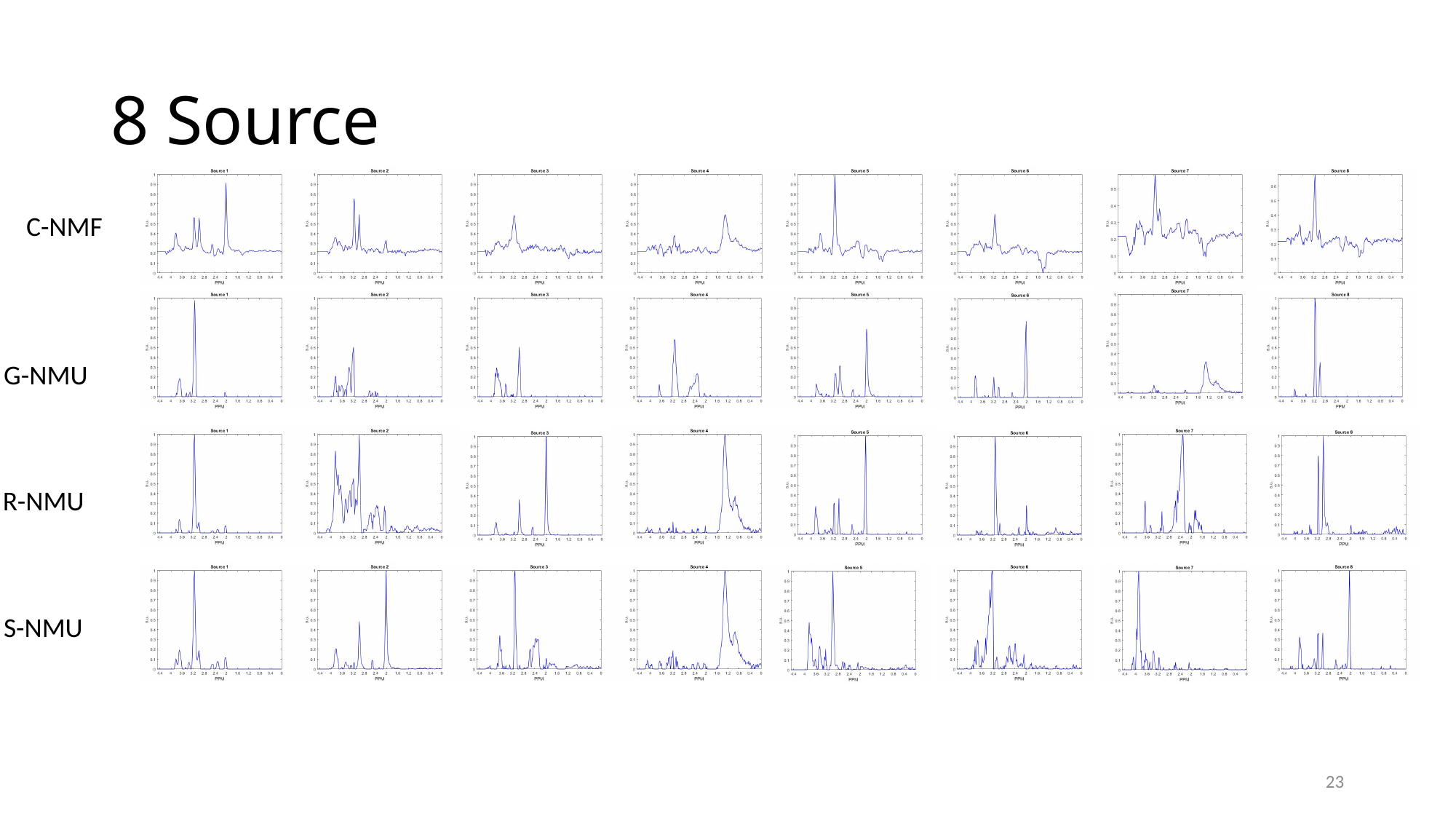

#### 8 Source
C-NMF
G-NMU
R-NMU
S-NMU
23

### Slide 24
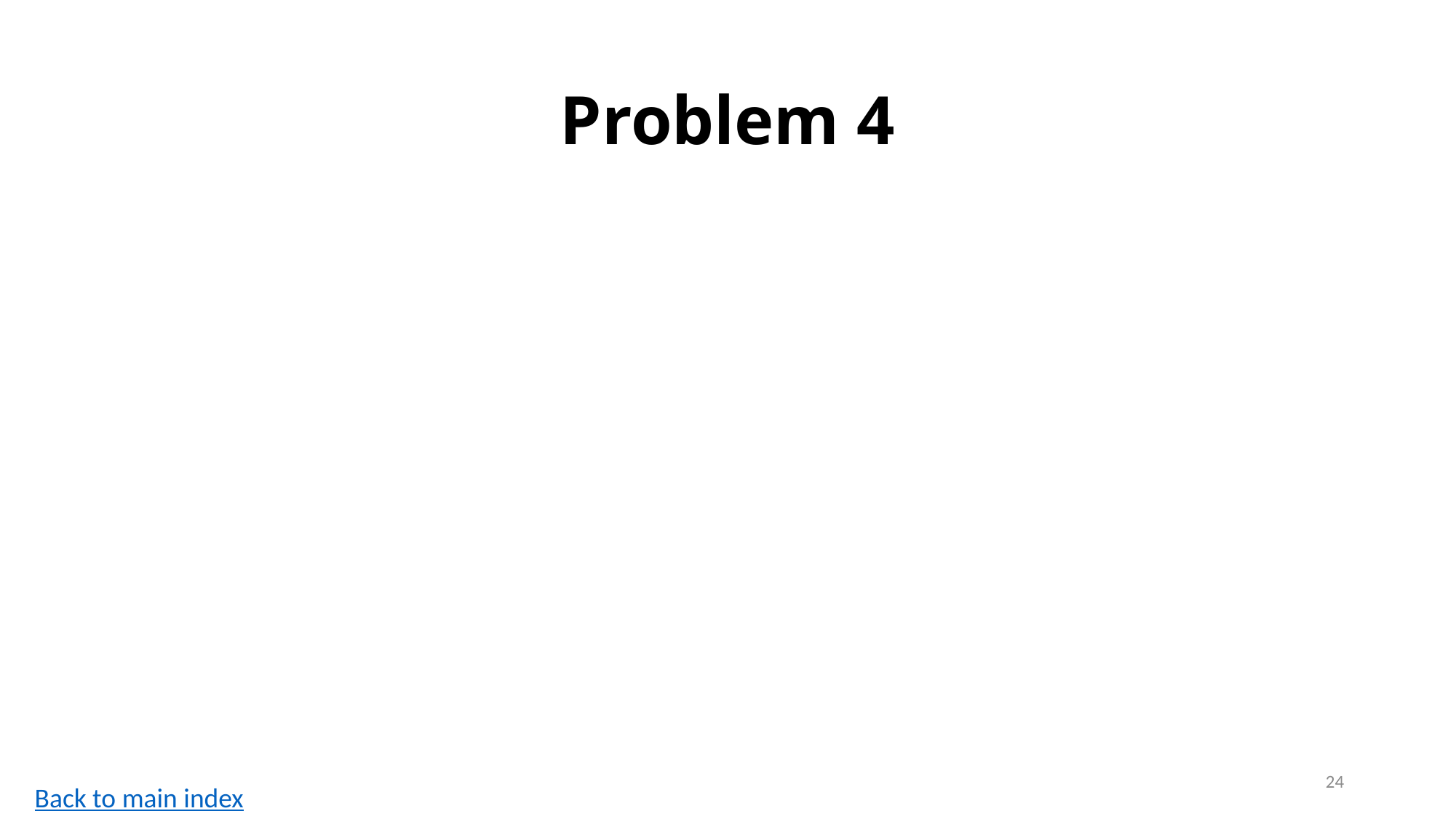

#### Problem 4
24
Back to main index

### Slide 25
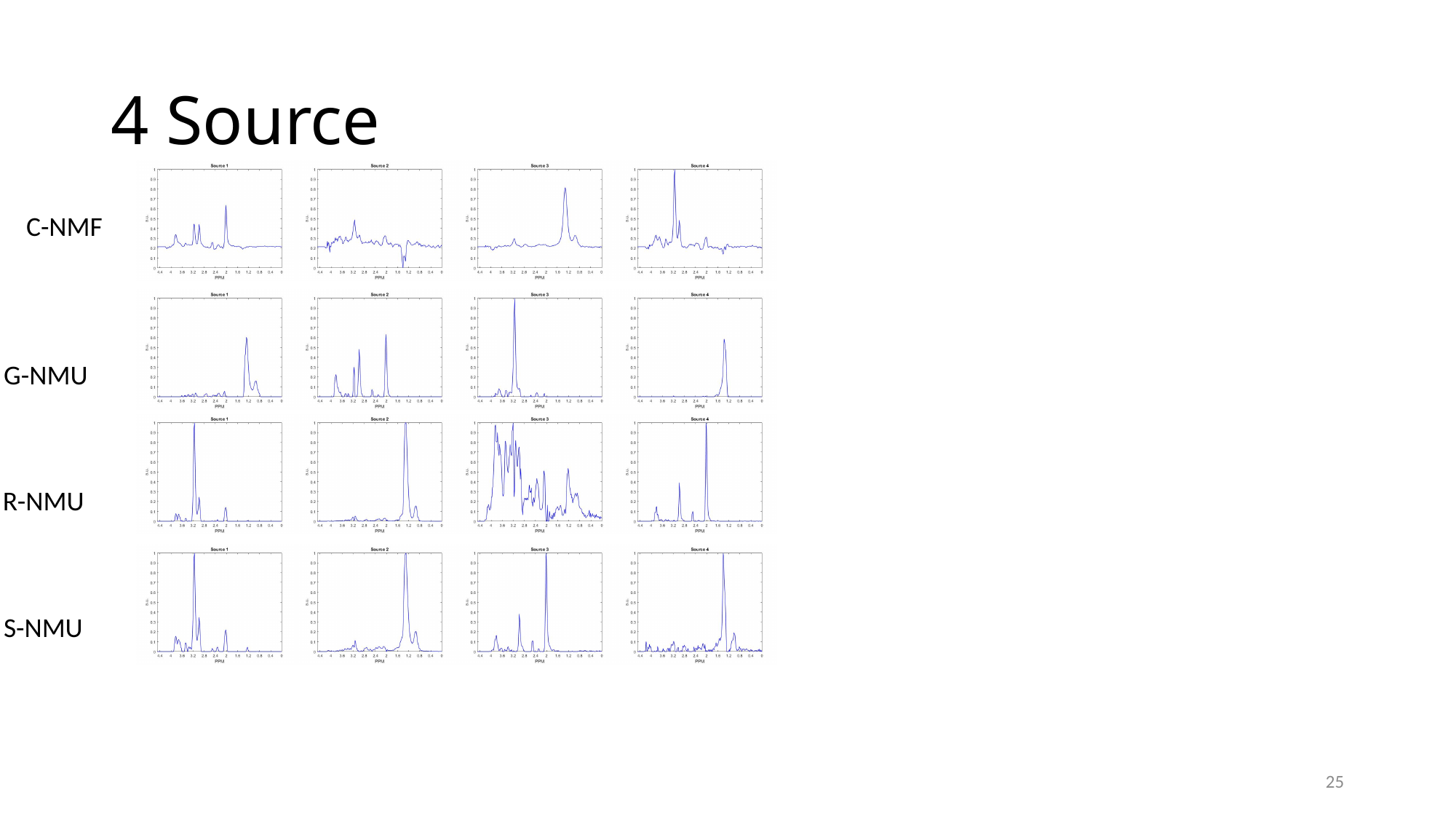

#### 4 Source
C-NMF
G-NMU
R-NMU
S-NMU
25

### Slide 26
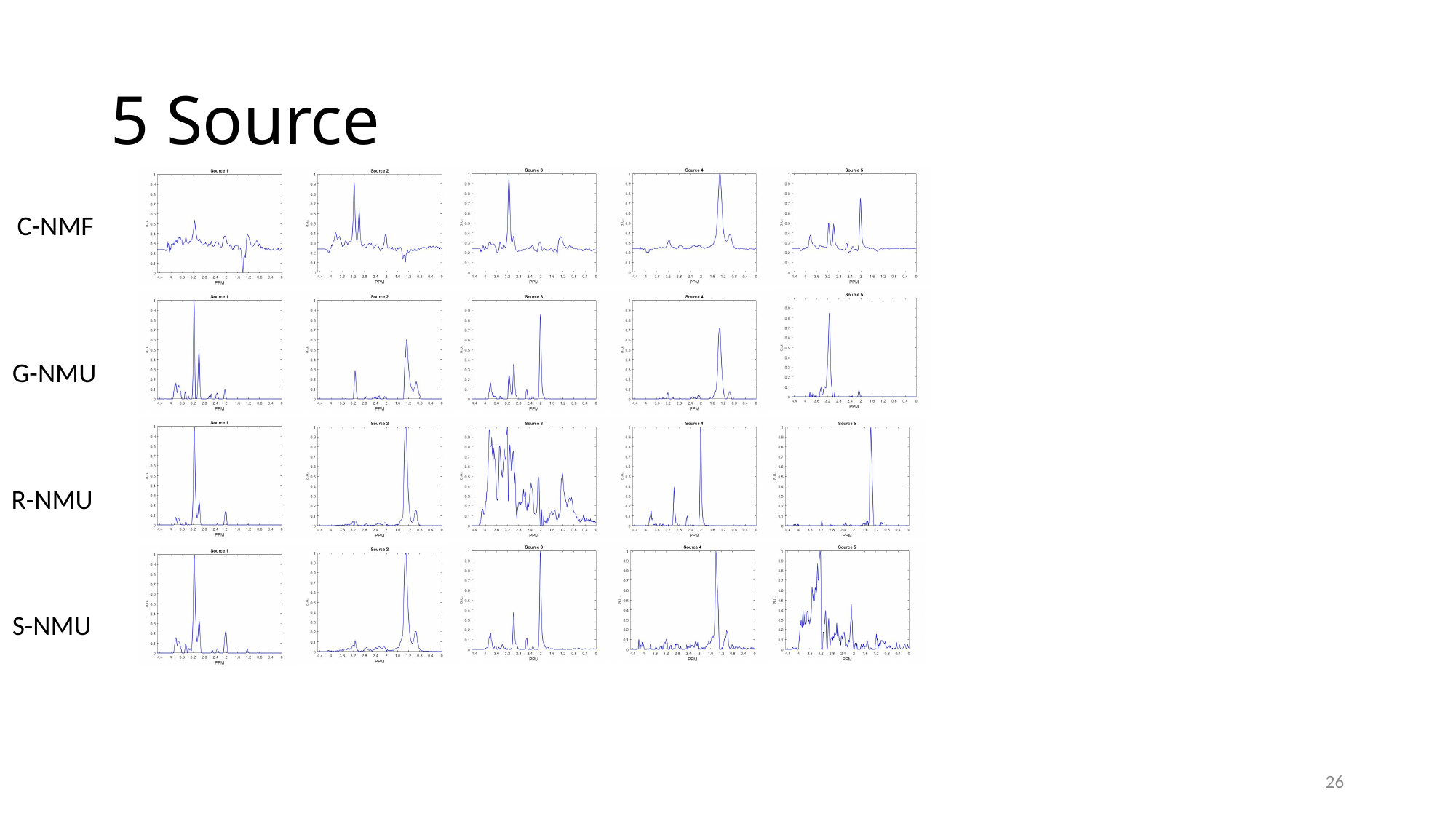

#### 5 Source
C-NMF
G-NMU
R-NMU
S-NMU
26

### Slide 27
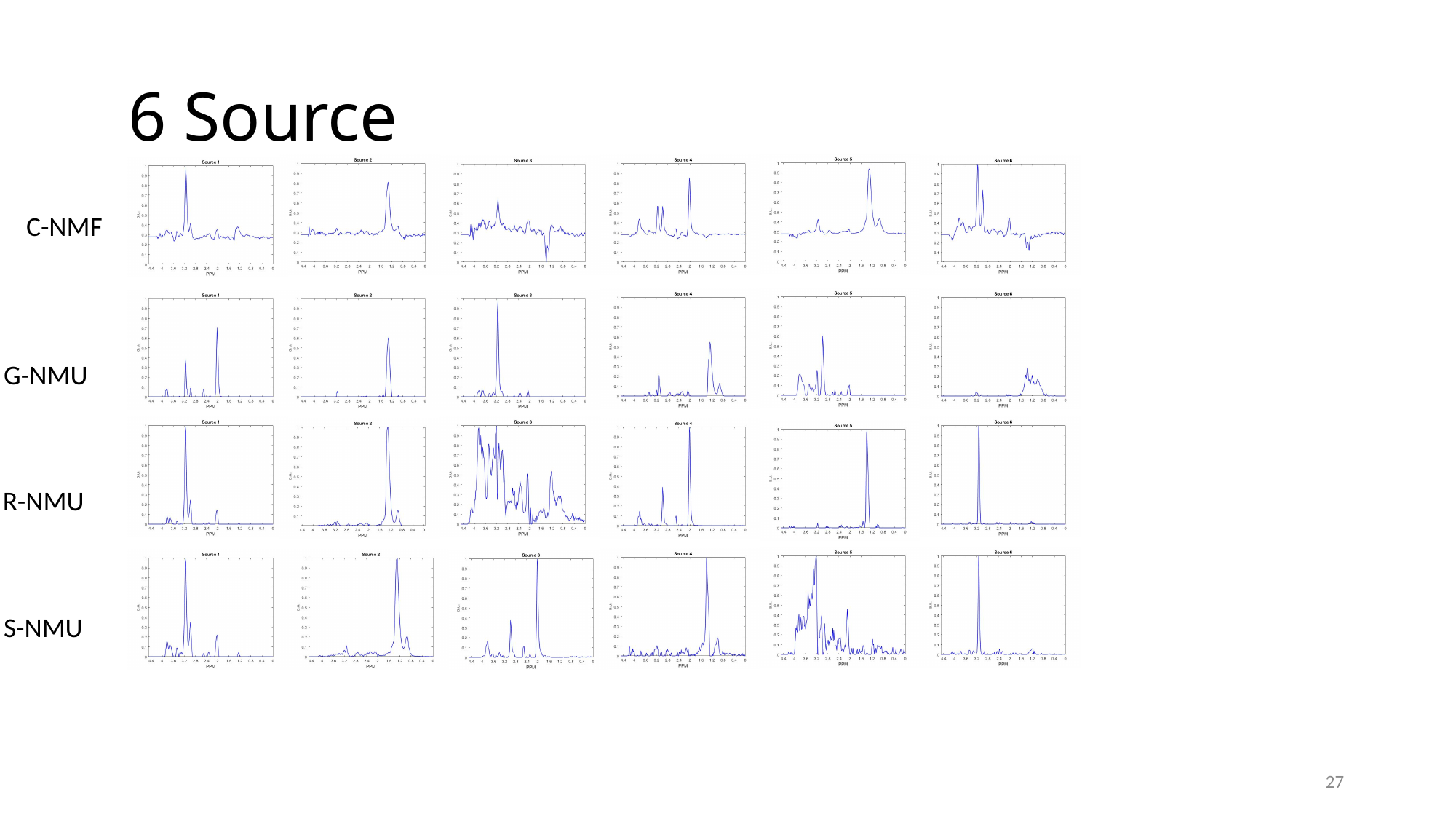

#### 6 Source
C-NMF
G-NMU
R-NMU
S-NMU
27

### Slide 28
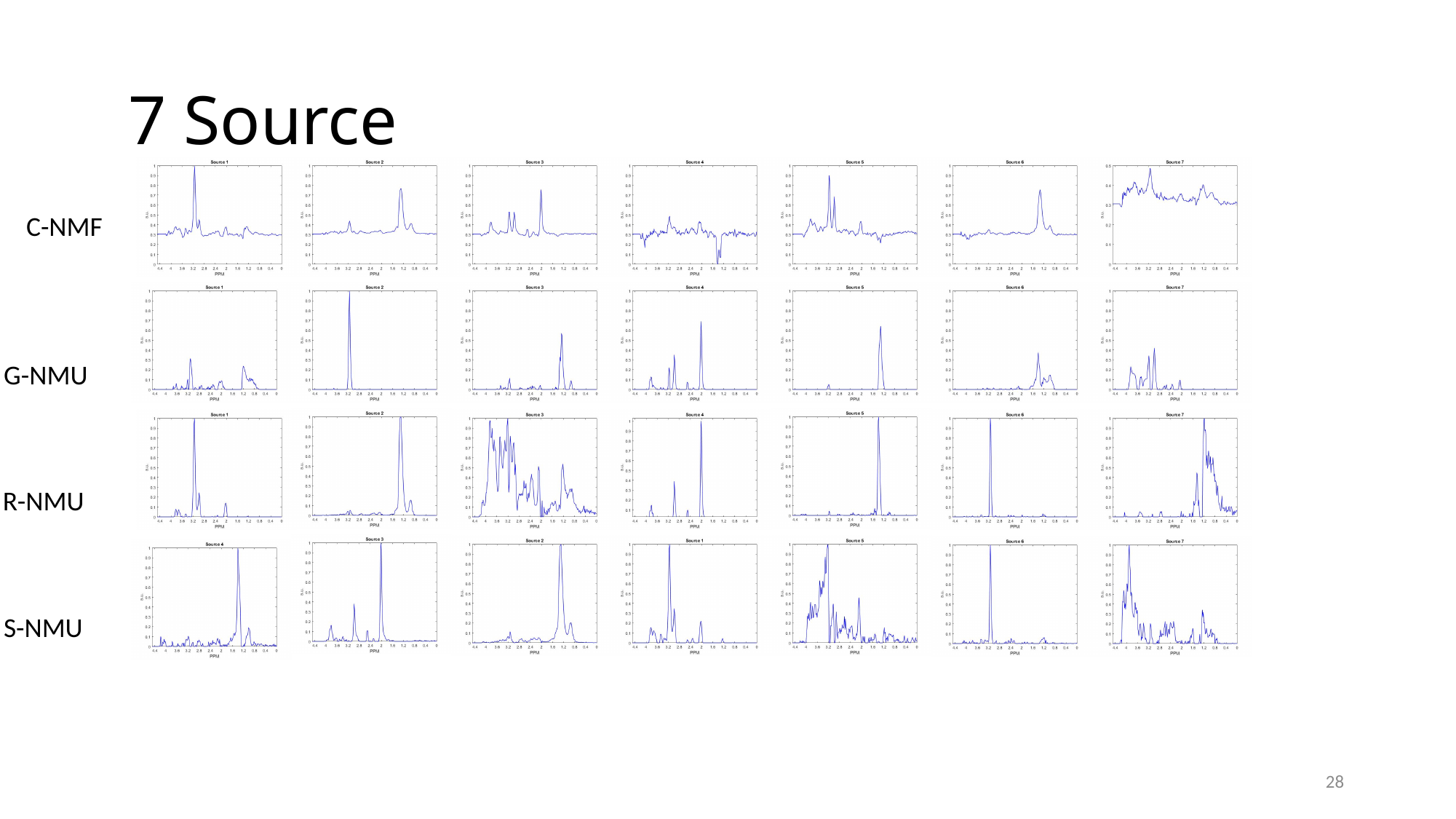

#### 7 Source
C-NMF
G-NMU
R-NMU
S-NMU
28

### Slide 29
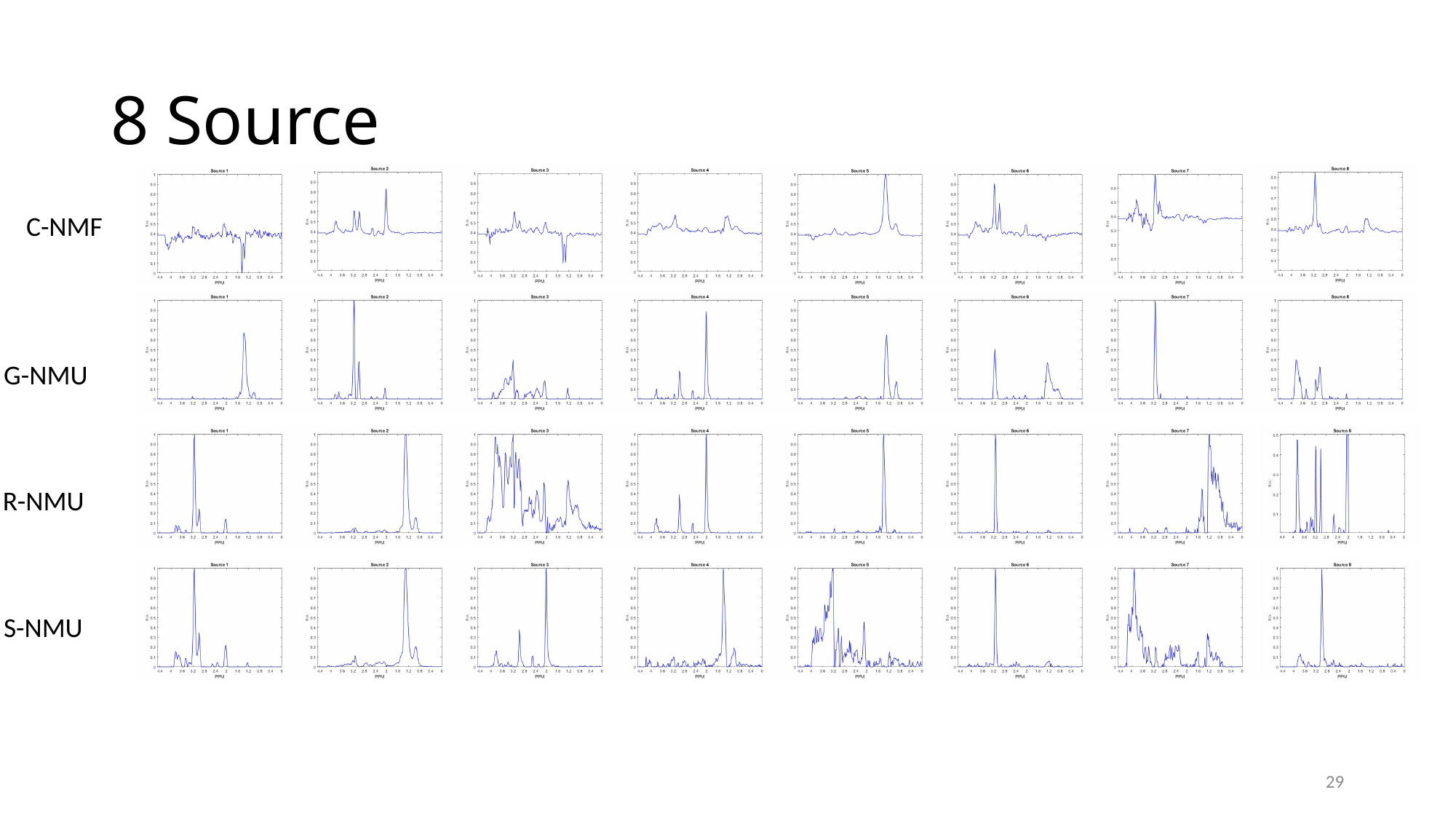

#### 8 Source
C-NMF
G-NMU
R-NMU
S-NMU
29

### Slide 30
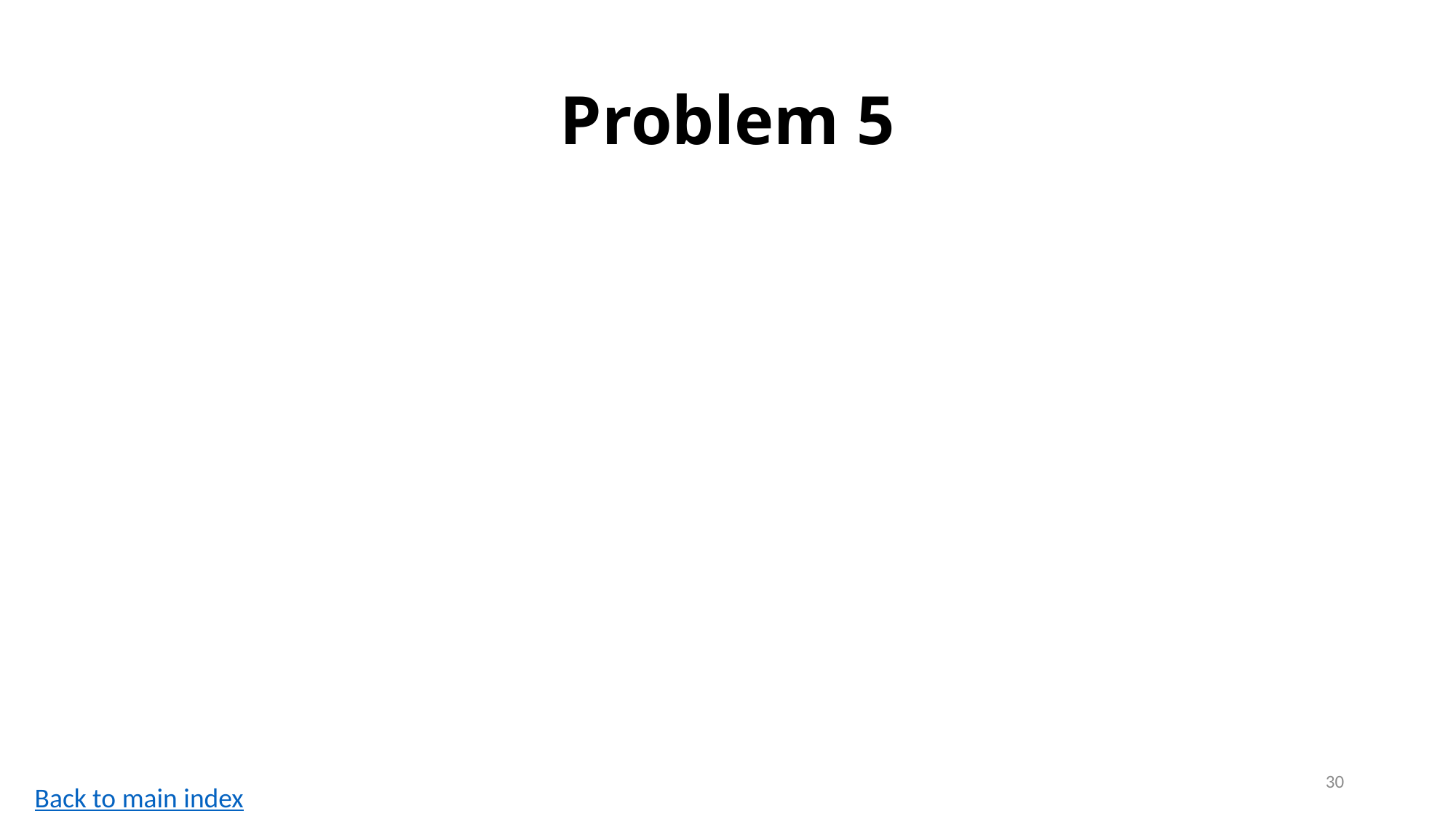

#### Problem 5
30
Back to main index

### Slide 31
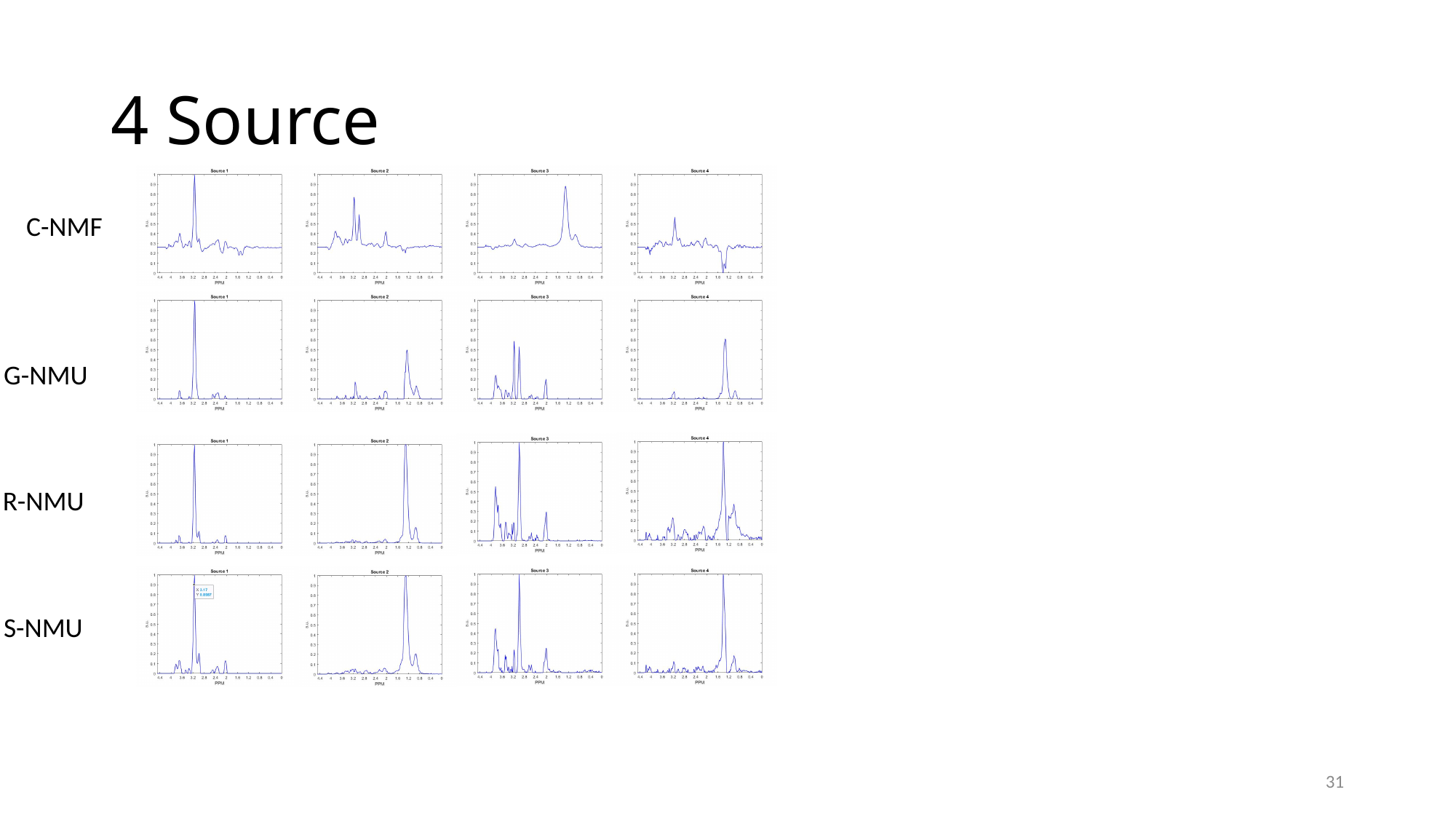

#### 4 Source
C-NMF
G-NMU
R-NMU
S-NMU
31

### Slide 32
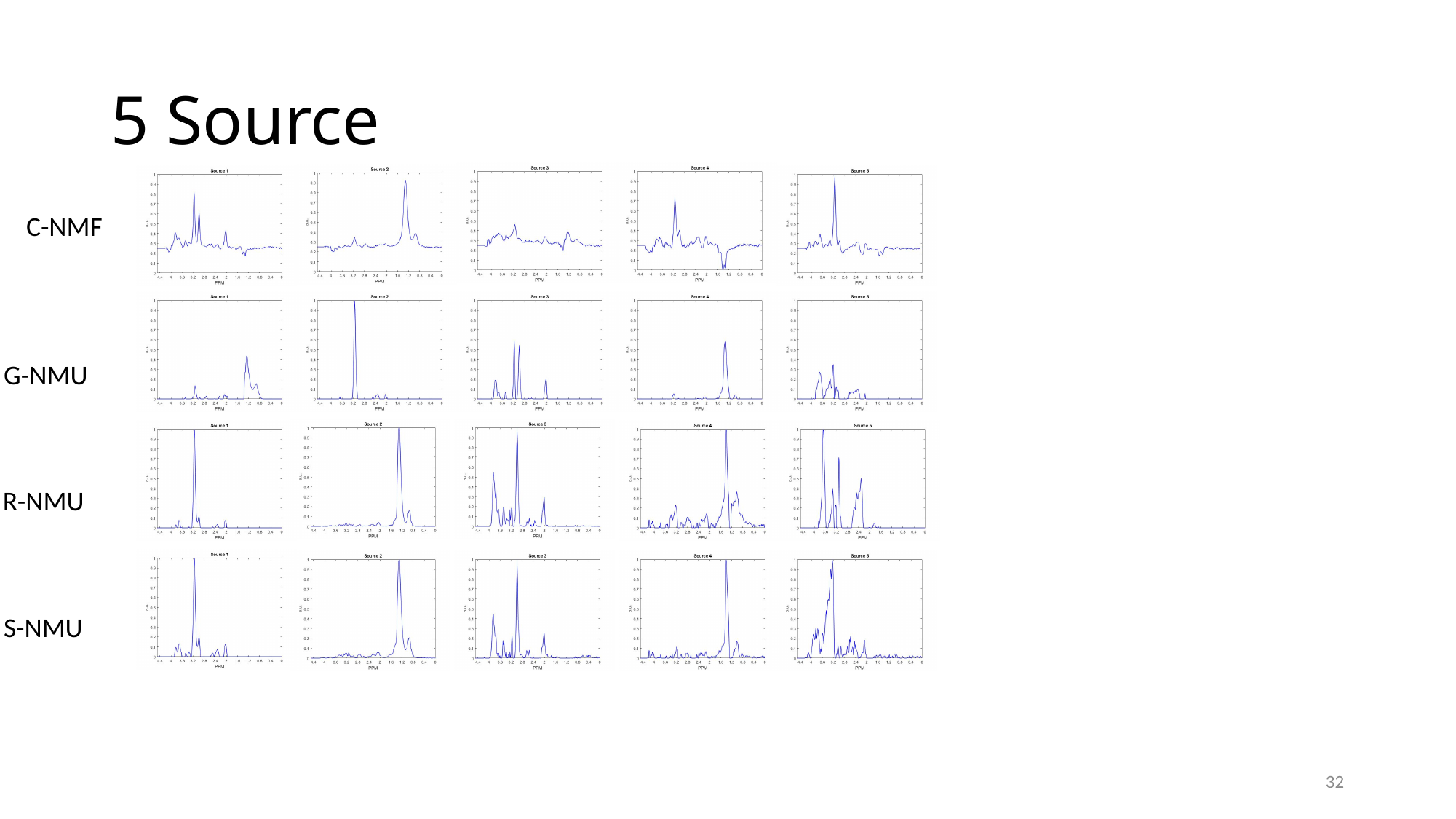

#### 5 Source
C-NMF
G-NMU
R-NMU
S-NMU
32

### Slide 33
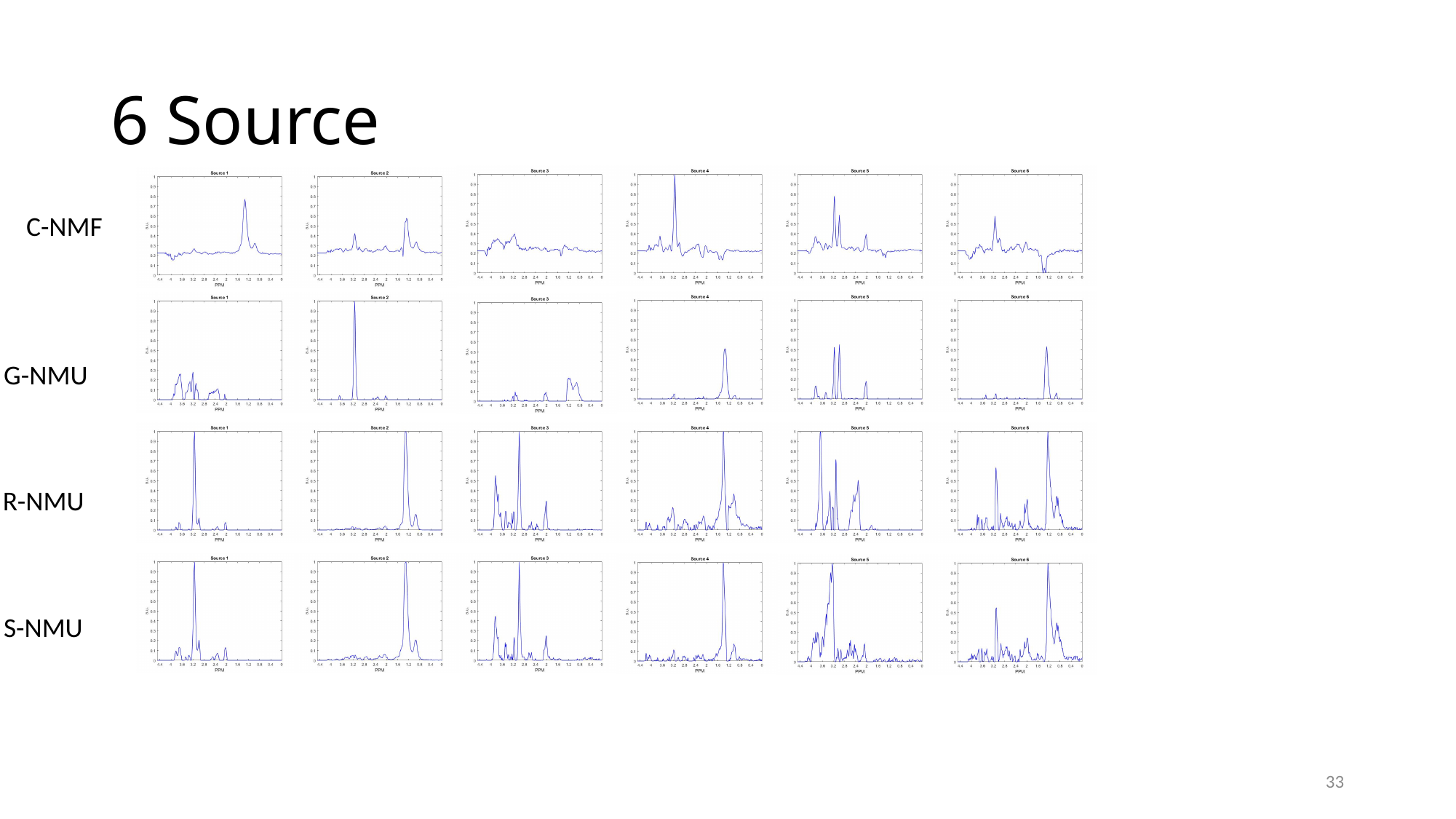

#### 6 Source
C-NMF
G-NMU
R-NMU
S-NMU
33

### Slide 34
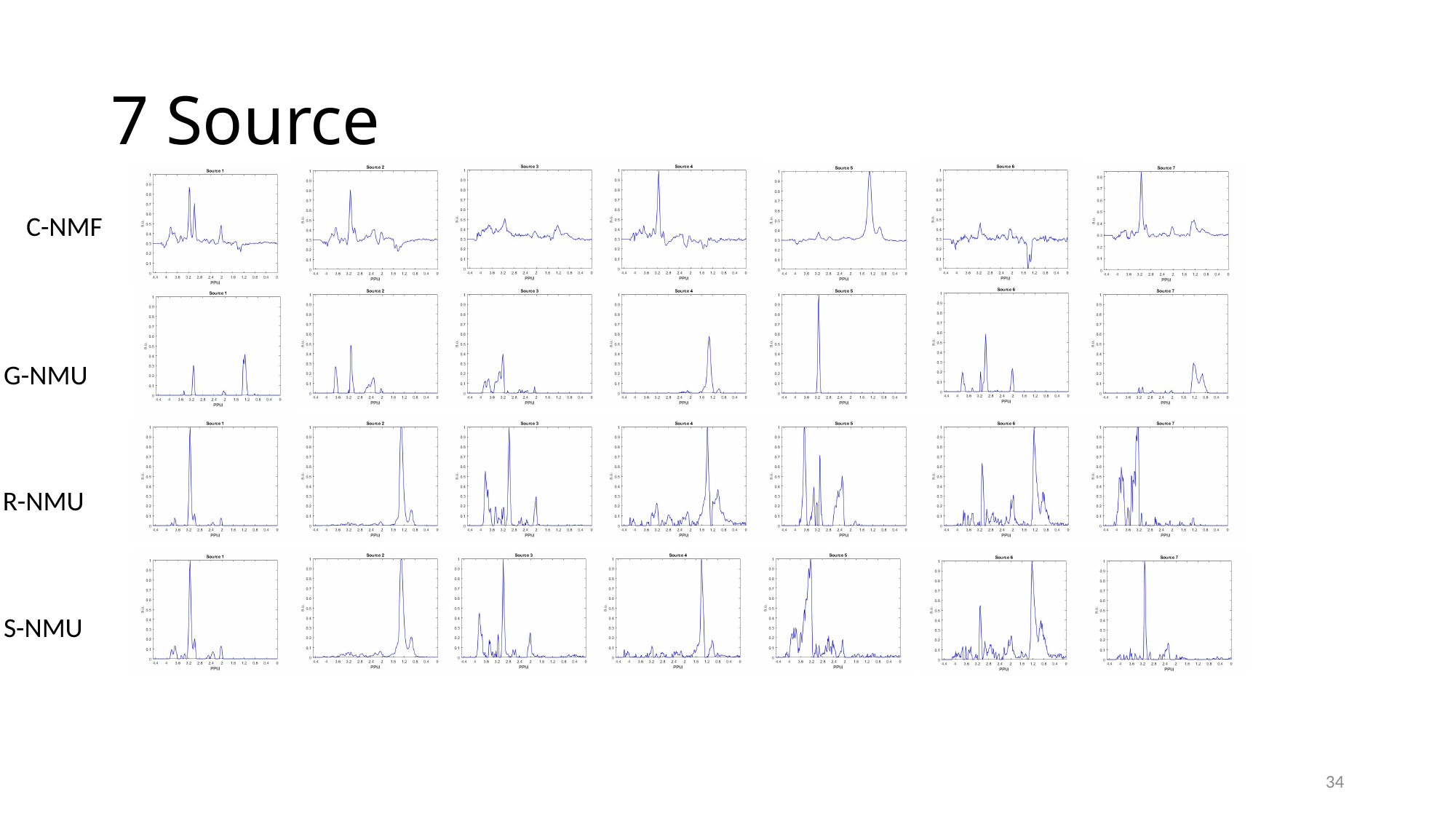

#### 7 Source
C-NMF
G-NMU
R-NMU
S-NMU
34

### Slide 35
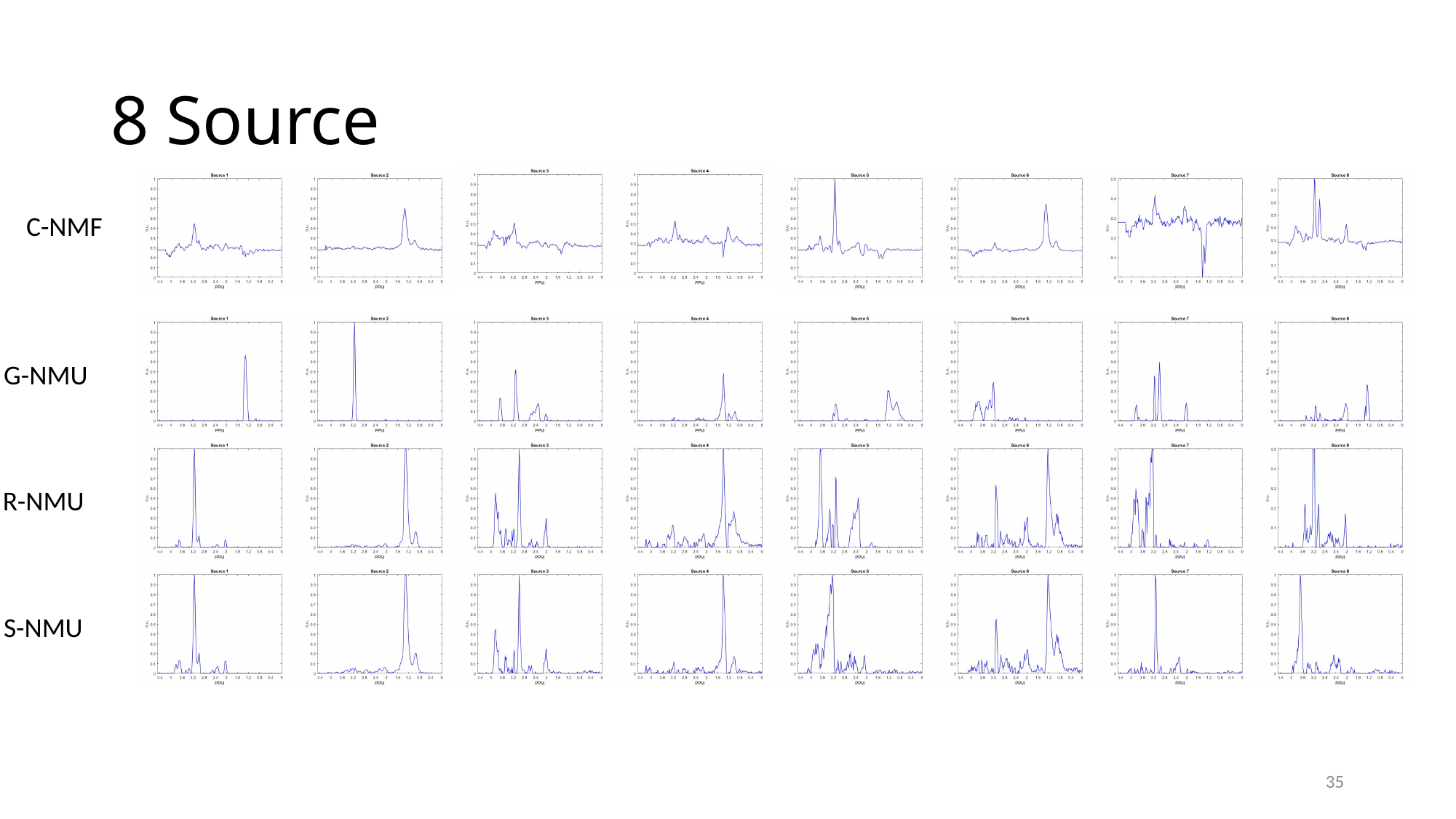

#### 8 Source
C-NMF
G-NMU
R-NMU
S-NMU
35
